# Dissecting depression symptoms: multi-omics clustering uncovers immune-related subgroups and cell-type specific dysregulation

**DOI:** 10.1101/2024.03.07.24303916

**Authors:** Jonas Hagenberg, BeCOME study group, OPTIMA study group, Tanja M. Brückl, Mira Erhart, Johannes Kopf-Beck, Maik Ködel, Ghalia Rehawi, Simone Röh-Karamihalev, Susann Sauer, Natan Yusupov, Monika Rex-Haffner, Victor I. Spoormaker, Philipp Sämann, Elisabeth Binder, Janine Knauer-Arloth

## Abstract

In a subset of patients with mental disorders, such as depression, low-grade inflammation and altered immune marker concentrations are observed. However, these immune alterations are often assessed by only one data type and small marker panels. Here, we used a transdiagnostic approach and combined data from two cohorts to define subgroups of depression symptoms across the diagnostic spectrum through a large-scale multi-omics clustering approach in 237 individuals. The method incorporated age, body mass index (BMI), 43 plasma immune markers and RNA-seq data from peripheral mononuclear blood cells (PBMCs). Our initial clustering revealed four clusters, including two immune-related depression symptom clusters characterized by elevated BMI, higher depression severity and elevated levels of immune markers such as interleukin-1 receptor antagonist (IL-1RA), C-reactive protein (CRP) and C-C motif chemokine 2 (CCL2 or MCP-1). In contrast, the RNA-seq data mostly differentiated a cluster with low depression severity, enriched in brain related gene sets. This cluster was also distinguished by electrocardiography data, while structural imaging data revealed differences in ventricle volumes across the clusters. Incorporating predicted cell type proportions into the clustering resulted in three clusters, with one showing elevated immune marker concentrations. The cell type proportion and genes related to cell types were most pronounced in an intermediate depression symptoms cluster, suggesting that RNA-seq and immune markers measure different aspects of immune dysregulation. Lastly, we found a dysregulation of the *SERPINF1*/VEGF-A pathway that was specific to dendritic cells by integrating immune marker and RNA-seq data. This shows the advantages of combining different data modalities and highlights possible markers for further stratification research of depression symptoms.

## 1. Introduction

Mental disorders significantly impact human health, health care systems, and economies, accounting for up to 16% of global disability-adjusted life years (Arias, Saxena, and Verguet 2022). Despite this substantial impact, the development of effective and innovative treatments remains a challenge (O’Donnell et al. 2019). Major depressive disorder (MDD) exemplifies these challenges, being the foremost psychiatric contributor to global disability (Vos et al. 2020).

The difficulty in understanding the underlying molecular causes of MDD arises from its biological heterogeneity. MDD manifests with a wide range of symptoms and is often comorbid with other mental disorders (Kessler et al. 2017; Kaufman and Charney 2000), which complicates its precise characterization. While the heritability of depression has been known for decades (McGuffin et al. 1996), genome-wide associations studies (GWASs) using minimal phenotyping have only yielded significant insights with extremely large sample sizes (Howard et al. 2019). Adopting a transdiagnostic approach that includes disorders across diagnostic categories and uses biological pathways, revealed associations between GWAS findings from MDD, schizophrenia and bipolar disorder with cytokine and immune pathways (O’Dushlaine et al. 2015).

Further evidence supports the pivotal role of the immune system in the pathology of MDD. Stress, a risk factor for depression, can lead to increased inflammation (Hammen 2015; Rohleder 2019). Accordingly, a subgroup of roughly 30% of MDD patients shows a proinflammatory profile - often referred to as low-grade inflammation - characterized by elevated levels of C-reactive protein (CRP) (Osimo et al. 2019). Our recent research corroborated this observation of immune-related depression, demonstrating genetic correlation between increased CRP level and depressive symptoms, e.g., tiredness, changes in appetite, anhedonia and feelings of inadequacy (Kappelmann et al. 2021).

Immune markers such as interleukin (IL)-6, IL-1beta and tumor necrosis factor (TNF) are often measured to evaluate immune dysregulation (Haapakoski et al. 2015) and contain genetic variants relevant for depression (Barnes, Mondelli, and Pariante 2017). Several studies have provided evidence that an inflammatory stimulus can lead to depressed mood, and that the pre-existing inflammatory status can predict the severity (Lasselin et al. 2020; Cho et al. 2019). For an overview on how cytokines and immune cells may mediate depressive symptoms see Miller and Raison 2016.

Many studies have employed case-control designs to find differences in immune markers between individuals with depression and healthy controls (Wittenberg et al. 2020; Sørensen et al. 2023; Sforzini et al. 2023). At the same time, various studies have established the link between specific depressive symptom profiles and inflammation. Increased inflammation has been observed in patients with dysregulated metabolism (immuno-metabolic depression) and linked to anhedonia (Lamers et al. 2013; Milaneschi et al. 2020; Felger et al. 2016; Lucido et al. 2021). A study by Franklyn et al. 2022 found that inflammation is associated with symptoms like altered eating patterns, appetite and tiredness. Lynall et al. 2020 focusing on immune cell counts, identified a subgroup of depressed patients with increased levels of monocytes, CD4+ T cells and neutrophils. This subgroup also demonstrated increased CRP and IL-6 levels, correlating with more severe depressive symptoms.

The potential of an inflammatory subtype goes beyond symptom profiles, as a study by Cattaneo et al. 2020 identified differences in whole blood gene expression related to inflammation that differentiate patients with treatment-resistant depression from those responding to treatment. Such findings highlight the potential of precisely characterizing immune dysfunction within MDD, aiming to identify patients who might benefit significantly from targeted immunomodulation. In recent years, several randomized control trials evaluated the effectiveness of adding an anti-inflammatory medication to antidepressants. Some of these trials adopted elevated CRP concentration as an inclusion criterion or a measure for secondary analyses. While there is some evidence for beneficial effects (Savitz et al. 2018; Köhler-Forsberg et al. 2019; Nettis et al. 2021), several other studies reported no discernible differences between patients treated with anti-inflammatory medication or those receiving placebos (Husain et al. 2020; Hellmann-Regen et al. 2022), even when stratified by CRP (Baune et al. 2021). Inspired by advancements in cancer research, Miller and Raison 2023 suggest shifting from traditional diagnostic evaluations to focusing on symptoms influenced by inflammation, such as anhedonia, changes in appetite and sleep (Drevets et al. 2022). This shift underpins our use of a transdiagnostic cohort, aiming to transcend diagnostic boundaries and explore common biological underpinnings across psychiatric conditions.

This push towards a more integrative and holistic understanding aligns with the broader field of psychoneuroimmunology. Traditionally, many studies in this area have relied on single omics approaches (e.g., transcriptomics or protein concentrations of peripheral immune markers). However, with the recent advancements in multi-omics techniques such as clustering or factor analysis methods (Li et al. 2021), there is an opportunity to integrate these individual omics data sets. Together with data from psychological assessments, imaging, and wearable devices (Moshe et al. 2021; Seppälä et al. 2019), we can further elucidate the complex interplay between the immune system and mental disorders by correlating molecular disease pathologies with clinical data (Mengelkoch et al. 2023; Halaris et al. 2019).

Building on this integrated approach, we selected a transdiagnostic sample of 237 individuals from two different cohorts with broad clinical assessments. Our goal was to comprehensively explore the relationship between the immune system and depression symptoms across the diagnostic spectrum utilizing multi-omics clustering techniques and considering a range of diagnoses of mental disorders. We hypothesized that distinct subgroups, defined by biological indicators of inflammation, would show different clinical and behavioral profiles. Furthermore, we measured an immune marker panel (p=43 markers) and the RNA-sequencing (seq) data (p=12210 genes) from their plasma and peripheral blood mononuclear cells (PBMCs), respectively. Using both the immune markers and RNA-seq data, we clustered the participants and characterized the unique clinical profiles of each subgroup (Figure 1).

**Figure 1:**
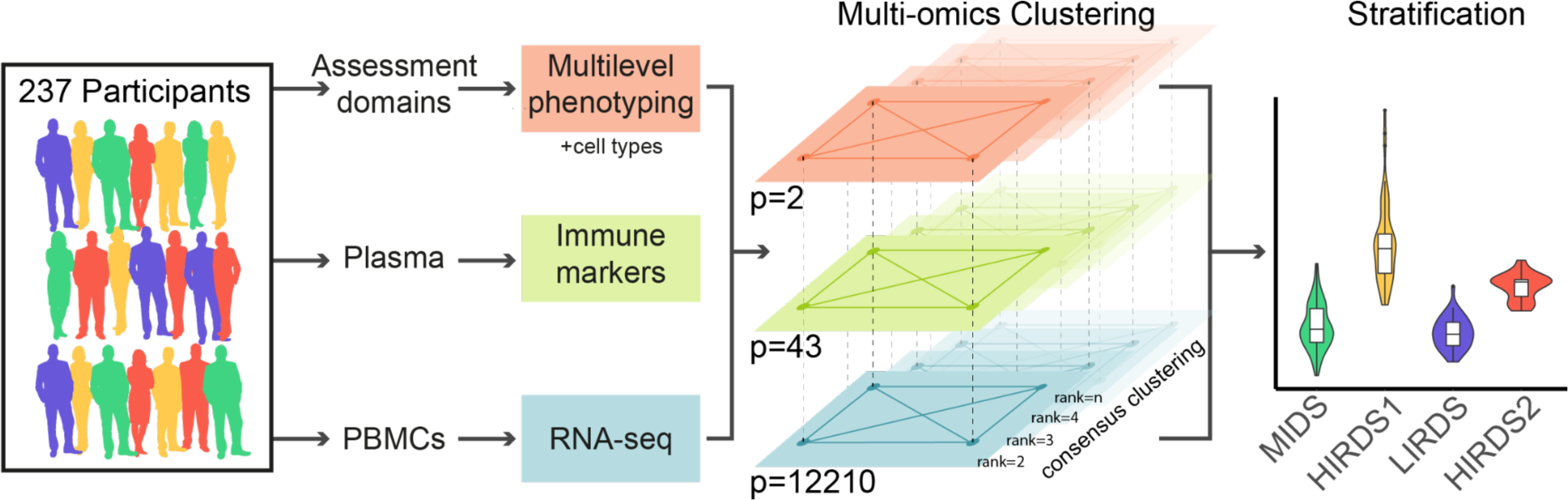
Analysis overview: 237 participants with a DSM-IV (subthreshold) diagnosis according to the Munich-Composite International Diagnostic Interview were clustered with a multi-omics clustering. The different layers contained phenotypes (age and BMI, orange), 43 immune markers measured in plasma (green) and RNA-seq from peripheral blood mononuclear cells (PBMCs, blue). The clustering was performed repeatedly as a consensus clustering on subsets of the data to obtain stability metrics for the optimal number of clusters. The stratification into clusters was characterized by several phenotypes. MIDS = mild depression symptoms cluster, HIRDS = high immune-related depression symptoms cluster, LIRDS = low immune-related depression symptoms cluster.

## 2. Methods

For detailed methods see the Supplementary Methods.

### 2.1 Sample selection

The study sample comprised 246 participants from the Biological Classification of Mental Disorders (BeCOME) study (ClinicalTrials.gov: NCT03984084, Bruckl et al. 2020) and 115 participants from the OPtimized Treatment Identification at the MAx Planck Institute (OPTIMA) study (ClinicalTrials.gov: NCT03287362, Kopf-Beck et al. 2020). The Munich-Composite International Diagnostic Interview (DIA-X/M-CIDI, Wittchen et al. 1995; Wittchen and Pfister 1997) was employed to assess all participants. Out of these with immune marker measurement, 237 affected participants (BeCOME=134, OPTIMA=103, Figure S1) met either threshold or subthreshold DSM-IV DIA-X/M-CIDI criteria for any substance use, affective or anxiety disorder including post-traumatic stress disorder and obsessive-compulsive disorder within the last 12 months. 192 of these participants had a (subthreshold) DSM-IV diagnosis of major depression or dysthymia. We also included 36 mentally healthy participants from the BeCOME study without any DSM-IV IDA-X/M-CIDI diagnosis as controls.

### 2.2 Ethics approval and informed consent

The BeCOME and OPTIMA studies were approved by the ethics committee of the Ludwig Maximilian University, in Munich, Germany, under the reference numbers 350–14 and 17– 395, respectively. Written informed consent was obtained from all participants before study enrollment.

### 2.3 Assessments

#### 2.3.1 Questionnaire data

All participants were assessed by the Beck Depression Inventory (BDI) II (Hautzinger, Keller, and Kühner 2006) and the Montgomery–Åsberg Depression Rating Scale (MADRS, Schmidtke et al. 1988). An overview about the sample characteristics for this subset are provided in Table 1, for more details see Table S1.

**Table 1:** Cohort overview with either percent of participants and absolute numbers in brackets or mean and standard deviation in brackets. The depression diagnosis was assessed by the Munich-Composite International Diagnostic Interview and includes full and subthreshold cases. Psychotropic drugs include antidepressants, moodstabilizer, neuroleptics, tranquilizer and herbal psychotropics. BDI = Beck Depression Inventory. Information on other diagnoses of mental disorders are available in Figure S5, on somatic diseases and medication in Table S1. MDD = major depressive disorder.

| status | n | age | female | BMI | MDD or dysthymia diagnosis | BDI | psychotropic drugs |
| --- | --- | --- | --- | --- | --- | --- | --- |
| case | 237 | 38.8<br>(13.3) | 58%<br>(138) | 25<br>(5.2) | 81% (192) | 23.6<br>(13.2) | 43% (103) |
| control | 36 | 31.4<br>(9.4) | 61%<br>(22) | 23.7<br>(2.8) | 0% (0) | 2 (2.9) | 0% (0) |

#### 2.3.2 Blood collection

Blood was collected in the morning under fasted conditions, peripheral blood mononuclear cells (PBMCs) were extracted and stored at the biobank.

#### 2.3.3 Immune marker measurements

We used the V-PLEX Human Biomarker 54-Plex Kit (Meso Scale Diagnostics (MSD), Rockville, USA, Cat. No. K15248G-2) to measure immune markers in plasma. In addition, enzyme-linked immunosorbent assay (ELISA) was used to measure the following markers: high-sensitivity C-reactive protein (hsCRP, Tecan Group Ltd., Männedorf, Switzerland, Cat. No. EU59151), cortisol (Tecan Group Ltd., Männedorf, Switzerland, Cat No. RE52061), interleukin (IL)-6 (Thermo Fisher Scientific, Waltham, USA, Cat. No. BMS213HS), IL-6 soluble receptor (sIL-6R, Thermo Fisher Scientific, Waltham, USA, Cat. No. BMS214) and IL-13 (Thermo Fisher Scientific, Waltham, USA, Cat. No. BMS231-3). All assays were performed according to the manufacturer’s instructions. Immune markers with high sensitivity alternatives or with more than 16% missing values, along with participants showing hsCRP levels suggestive of acute infection, were excluded. This resulted in 43 markers and 273 participants remaining for analysis (Table S2 and Figure S1).

#### 2.3.4 RNA extraction and sequencing

The RNA was extracted with the chemagic 360 instrument (PerkinElmer, Waltham, USA), rRNA depleted, libraries prepared with the Lexogen CORALL total RNA-Seq Library Prep Kit with UDIs 12 nt Sets A1-A4 (Cat. No. 117.96, 132.96, 133.96, 134.96) and sequenced on an NovaSeq 6000 (Illumina, San Diego, USA) yielding an average of 30.6 million paired reads per library.

#### 2.3.5 Structural magnetic resonance imaging (MRI) data assessment

151 affected participants (BeCOME=117 and OPTIMA=34) and 29 healthy controls had high resolution T1-weighted images with the identical sequence in both original studies (Sagittal FSPGR 3D BRAVO, TE 2.3 ms, TR 6.2 ms, TI 450 ms, FA 12°, FOV 25.6 × 25.6 × 20.0 cm3, matrix 256 × 256, voxel size 1 × 1 × 1 mm3) available.

#### 2.3.6 Electrocardiography data assessment

149 affected participants (BeCOME=108 and OPTIMA=41) and 32 healthy controls had an electrocardiography (ECG) recording that spanned 24h including a sleeping period with the portable ECG-device Faros 180 (Mega Electronics Ltd, Kuopio, Finland) at a sampling frequency of 500 Hz available.

### 2.4 Data analysis

#### 2.4.1 Immune marker analysis

The immune markers were quantile-normalized and corrected for the biobank storage position. For each immune marker, a linear model was fitted in R version 4.0.2 (R Core Team 2023) to regress out the batch variable. The residuals from this regression were then used for downstream analysis.

#### 2.4.2 RNA-seq analysis

The reads were aligned to GRCh38.p12 (Ensembl version 97, Martin et al. 2023) with STAR version 2.7.7a (Dobin et al. 2013) and counted with featureCounts version 1.6.4 (Liao, Smyth, and Shi 2014).

Quality control and exclusion of genes with few counts or high GC content influence left 229 affected participants, 33 controls and 12210 genes (Figure S1). The data was sequentially corrected for the GC content and a preparation batch variable with ComBat_seq from the sva package version 3.38.0 (Leek et al. 2021) and normalized with vst from DESeq2 version 1.30.1 (Love et al. 2021).

#### 2.4.3 Cell type deconvolution

The filtered RNA-seq count data, which was not batch-corrected, was converted to TPM and deconvoluted using the granulator package version 1.6.0 (Pfister, Kuettel, and Ferrero 2023) in R version 4.2.0 (R Core Team 2023). All cell types with a standard variation of less than 0.5 were excluded, resulting in 14 cell types.

#### 2.4.4 Multi-omics clustering and statistical analysis of cluster differences

Each variable of the normalized and batch corrected immune marker and RNA-seq data as well as age and BMI was standardized, the distance between the participants calculated and the clustering method SUMO version 0.3.0 (Sienkiewicz et al. 2022) was applied repeatedly on a subsample to calculate stability metrics for selecting the number of clusters.

If not otherwise stated, we used R version 4.0.2 (R Core Team 2023) and determined the variable importance based on the F-value from ANOVA tests. A higher variable importance indicates a more distinct separation between the clusters. For continuous variables not used in the clustering, we applied the Tukey Honest Significant Differences (TukeyHSD) test and reported adjusted p-values, for categorical variables the Fisher’s exact test.

We did not report p-values for group differences for variables that were used in the clustering as differences were anticipated (Kriegeskorte et al. 2009).

#### 2.4.5 Gene set analysis

For each participant, a score was calculated for every biological process and molecular function GO term gene set using GSVA version 1.36.3. (Hänzelmann, Castelo, and Guinney 2013) based on the batch-corrected and normalized RNA-seq data. The distribution of these gene set scores across clusters was tested using an ANOVA and F-values reported as variable importance.

#### 2.4.6 Prediction of responder status

The baseline values from gene expression data with accession number GSE45468 were mapped to ENSEMBL IDs. 8431 genes overlapped with our data and were used in a lasso model using glmnet version 4.1-8 (Friedman et al. 2023), the p-value was empirically calculated based on random genes.

#### 2.4.7 Imaging analysis

Morphological differences were investigated at the global, regional (FreeSurfer) and voxel level. FreeSurfer v7.1.1 (Fischl 2012) based phenotypes were restricted to those with strong meta-analytical evidence for an effect in MDD (Schmaal et al. 2016; 2017). Analysis of covariance (ANCOVA) was applied to the clusters covarying for intracranial volume, age and sex, and cluster effects were FDR corrected at 5%.

#### 2.4.8 Electrocardiography analysis

Raw data was preprocessed using the PhysioNet Cardiovascular Signal Toolbox (Vest et al. 2018) in Matlab version 2020b (The MathWorks Inc. 2020). Time domain metrics were calculated for every minute using a sliding window of 5 minutes.

#### 2.4.9 Single-cell RNA-seq data analysis

Single cell PBMC RNA-seq data from 14 individuals with depression and other mental disorders with accession number GSE185714 was processed with scanpy version 1.9.3 (Wolf, Angerer, and Theis 2018) according to Schmid et al. 2021 and their cell type labels used.

## 3. Results

### 3.1 Multi-omics clustering with age, BMI, immune markers and RNA-seq data identified 4 clusters with different depression severity and inflammation status

To discover subgroups with immune-related depression symptoms, we assessed a comprehensive immune marker panel (p = 43 markers) along with matched RNA-seq data (p = 12210 genes) in 237 participants (Figure S1) diagnosed with MDD or another stress-related mental disorder within the last 12 months. The mean age of the affected participants was 39 years and the mean BMI 25 (see Table 1 for sample characteristics). Besides the biological data, the participants underwent phenotypic assessment. Our approach involved clustering the participants based on multiple biological datasets and subsequently characterizing these subgroups using psychometric data.

For multi-omics clustering, we employed SUMO (Sienkiewicz et al. 2022), a non-negative matrix factorization based clustering algorithm, that provides cluster stability metrics via consensus clustering. This method organizes variables into layers and computes the distance between the participants within each layer, which subsequently informs the clustering process. For our study, age and BMI were combined as one layer while the immune markers and RNA-seq data were each defined as separate layers. According to the built-in stability metrics (Figure S2A-B), this approach yielded a four cluster solution. These clusters included 121, 58, 39 and 19 participants, respectively (Table S3). In light of the findings described below, we termed cluster 1 *mild depression symptoms* cluster (MIDS), cluster 2 *high immune-related depression symptoms* cluster (HIRDS) 1, cluster 3 *low immune-related depression symptoms* cluster (LIRDS) and cluster 4 HIRDS2. Each of the identified clusters showed a different pattern of immune markers and contained participants from both the BeCOME and OPTIMA cohorts, which differ in the in- and outpatient ratio, psychopharmacological drug use and sex distribution (see Figures 2A and S3). The significantly different distribution of the cohorts across the clusters (Fisher’s exact test, p = 0.000003) was driven by the MIDS cluster; between the LIRDS and HIRDS clusters there was no different distribution (Fisher’s exact test, p = 0.507320).

**Figure 2:**
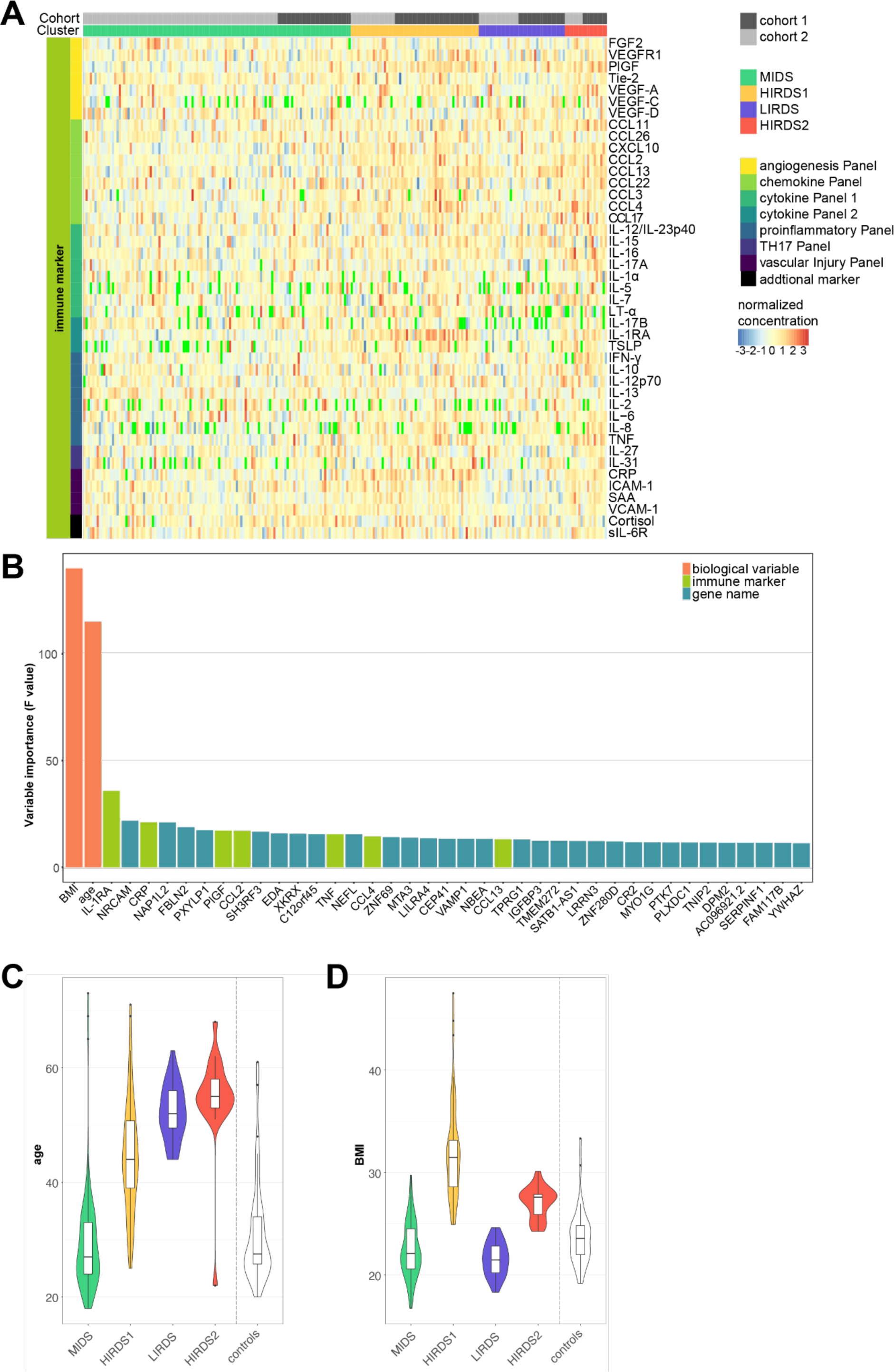
Initial clustering with age, BMI, immune markers and RNA-seq data. (A) Heatmap of immune markers where the columns are participants while the cohort and cluster are noted at the top. The color denotes the normalized concentration with green values missing. (B) Barplot displaying the variable importance of the top 40 variables that most differentiate the clusters, calculated using the F-value from an ANOVA model with the clusters as independent variables. These variable importance values should not be interpreted as p-values, because the variables were used in the clustering themselves and therefore inflate the p-values. Violin plots depicting the distribution of (C) age and (D) BMI across the clusters. N = 36 healthy controls not used in the clustering, MIDS = mild depression symptoms cluster, HIRDS = high immune-related depression symptoms cluster, LIRDS = low immune-related depression symptoms cluster, cohort 1 = OPTIMA, cohort 2 = BeCOME.

#### 3.1.1 BMI, age, IL-1RA, *NRCAM* and CRP identified as top differentiating variables

In order to understand which variables drive the clustering, we calculated the variable importance (VI) based on an ANOVA model (Table S4). Figure 2B illustrates that BMI (VI = 139.8) and age (VI = 114.8) were most important for the clustering, followed by interleukin 1 receptor antagonist (IL-1RA, VI = 35.7). Other important immune markers included high-sensitivity C-reactive protein (CRP, VI = 21), placental growth factor (PlGF, VI = 17.1), C-C motif chemokine 2 (CCL2 or MCP-1, VI = 17), tumor necrosis factor (TNF, VI = 15.4), CCL4 (MIP-1beta, VI = 14.4) and CCL13 (MCP-4, VI = 13.1). Among the genes, neuronal cell adhesion molecule (*NRCAM*, VI = 21.8), nucleosome assembly protein 1 like 2 (*NAP1L2*, VI = 21), fibulin 2 (*FBLN2*, VI = 18.7), 2-phosphoxylose phosphatase 1 (*PXYLP1*, VI = 17.4), SH3 domain containing ring finger 3 (*SH3RF3*, VI = 16.6), ectodysplasin A (*EDA*, VI = 15.8) and XK related X-linked (*XKRX*, VI = 15.7) were the most important. The genetic location of these genes are noted in Table S5, compared to the overall chromosomal distribution the top 100 genes show an enrichment in chromosome 7 (Figure S4). The importance of age and BMI in the clustering process was reflected in their distribution across clusters, as shown in Figure 2C-D. Age demonstrated an increase from MIDS through to HIRDS2. Notably, HIRDS2 and especially HIRDS1 showed elevated BMIs (mean BMI of 27 and 32, respectively) compared to the other clusters.

Next, we aimed to provide insights into the distribution of age, BMI, and CRP concentration across the clusters due to their importance in the clustering and the established role of CRP as a standard marker for inflammation. Notably, in all clusters except HIRDS2, a positive correlation was observed between CRP concentration and BMI. Specifically, for the MIDS cluster the Pearson correlation was 0.13 (t-test, p = 0.155); for HIRDS1, it was 0.40 (t-test, p = 0.002); for LIRDS, it was 0.39 (t-test, p = 0.015); and for HIRDS2 it was −0.61 (t-test, p = 0.006), as illustrated in Figure 3A. It also shows that the HIRDS clusters predominantly contained individuals with elevated CRP concentration and elevated BMI. As illustrated in Figure 3B, no significant correlation between age and CRP was discernible within the clusters, with Pearson correlations ranging between −0.16 and 0.01. Interestingly, the LIRDS cluster contained individuals with a restricted age range. Further examination revealed no strong correlation between age and BMI except for HIRDS2, with Pearson correlations ranging from 0.09 to 0.40, which were not significant (Figure 3C). Notably, while the individuals with a high BMI in HIRDS1 spanned a wide age range, HIRDS2 consisted mostly of individuals aged 50 or above.

**Figure 3:**
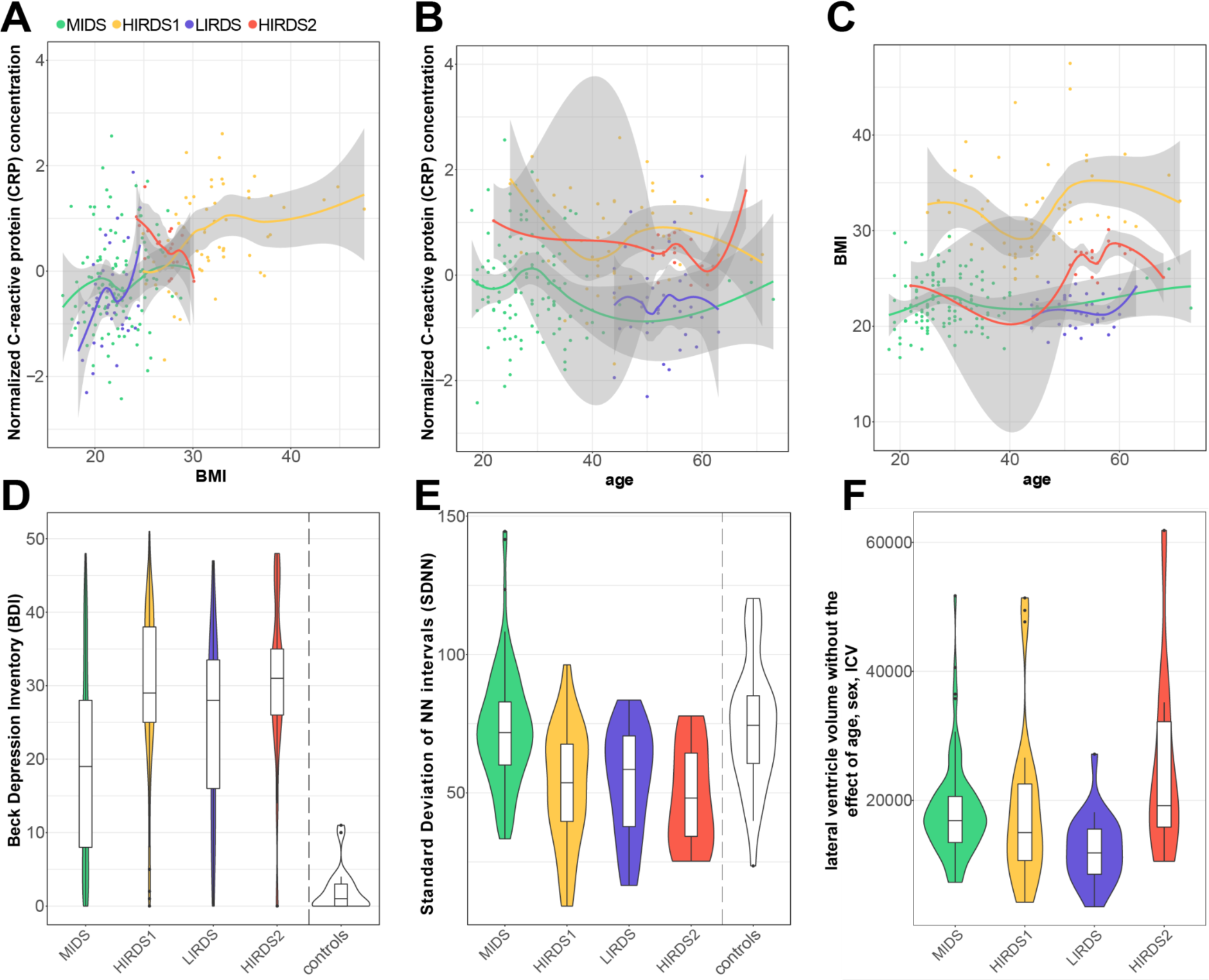
Distribution of biological and phenotype data across the clusters in the initial clustering with age, BMI, immune markers and RNA-seq data. (A) - (C) Scatterplots illustrate the associations between C-reactive protein (CRP), BMI and age across the four clusters, as determined by locally estimated scatterplot smoothing (LOESS). (D) Violin plots showing the distribution of Beck Depression Inventory (BDI)-II across the clusters. Significant differences (p < 0.05) were observed when comparing HIRDS1, LIRDS and HIRDS2 to the MIDS cluster. (E) Violin plots illustrating the Standard Deviation of normal-to-normal intervals (SDNN), a measure for heart rate variability, for 149 participants. The significant differences (p < 0.05) were observed between HIRDS1, LIRDS and HIRDS2 compared to the MIDS cluster. (F) Violin plots illustrating the the cluster-specific effect (global intercept + cluster-specific effect + residuals) of the lateral ventricle volume from a model including age, sex and intracranial volume (ICV) derived from structural magnetic resonance imaging for 151 participants. A significant difference (p < 0.05) was observed between the clusters. Because of the modeling approach, no value for the controls can be shown. MIDS = mild depression symptoms cluster, HIRDS = high immune-related depression symptoms cluster, LIRDS = low immune-related depression symptoms cluster.

#### 3.1.2 Clinical phenotypes, imaging features and heart rate data identified clusters with elevated depression severity

Next, we explored how the clusters, which were defined solely based on biological data (including immune markers, RNA-seq data, BMI, and age) without considering depression severity or symptoms, corresponded to these measures of depression post-clustering.

Interestingly, the LIRDS and HIRDS clusters exhibited greater depression severity, as measured by the Beck Depression Inventory-II (BDI), than the MIDS cluster (Tukey HSD test with adjusted p values, p = 0.028961, p = 0.000003 and p = 0.001172, respectively). This severity was especially pronounced in the HIRDS clusters (Figure 3D). A similar result was observed using the Montgomery–Åsberg Depression Rating Scale (MADRS) with p-values of 0.009829, 0.000466 and 0.003718 (Tukey HSD test with adjusted p values) for the LIRDS and HIRDS clusters, respectively. The proportion of individuals with either no depression or dysthymia diagnosis was not equal across the clusters (Fisher’s exact test, p = 0.000500, Figure S5), the MIDS cluster had the highest proportion of participants without such a diagnosis and also the lowest depression severity. While no single item of the BDI differentiated the cluster, there was a trend towards increased sleeping and appetite in the HIRDS clusters as indicated by the BDI (Figure S6A-B).

To further assess if the identified symptom differences were mirrored in other biological data, we analyzed heart rate data collected from portable ECG devices (n = 149) and structural imaging-derived features (n = 151) in a subset of the participants for whom these data were available. While there were no discernable differences in the median minimum or maximum normal-to-normal intervals between heartbeats between the clusters (Figure S6C), the heart rate variability metrics - for short-term (RMSSD) and long-term (SDNN) - were lower in the LIRDS and HIRDS clusters in comparison to the MIDS cluster (Figure 3E). The p-values for these differences ranged from 0.000039 to 0.005874 for RMSSD and from 0.000112 to 0.005446 for SDNN (Tukey HSD test with adjusted p values). The clusters showed a trend of different total brain volume (TBV) (F = 2.406, p = 0.070), with LIRDS showing higher TBV compared to the MIDS and HIRDS clusters; a similar pattern was seen for gray matter (GM) and white matter (WM) (Table S6). Regional analyses of 17 MDD-related phenotypes revealed robust effects on the lateral ventricle volume (F = 5.398, FDR adjusted p = 0.029, with the lowest volume in LIRDS, see Figure 3F), and nominal significant effects for the right rostral anterior cingulate and the bilateral hippocampus (Table S7). Voxel-based morphometry revealed no cluster main effects.

#### 3.1.3 Immune markers and genes revealed differences in immune system and brain related pathways between clusters

We wanted to understand how the immune markers and RNA-seq data each contributed to defining the clusters, and if these different data types contained distinct information.

Interestingly, except for CCL13, all the top important immune markers were elevated in the HIRDS clusters (Figure 4A). These clusters not only showed increased BMI but also the highest BDI. This distinctive pattern was reflected in CCL13’s low correlation with key markers such as IL-1RA and CRP (Figure 4D). While still discriminating the HIRDS clusters from the others, PlGF also had a similar low correlation with the top markers IL-1RA and CRP (Figure 4D). All top immune markers showed a positive correlation with BMI (Figure S7).

**Figure 4:**
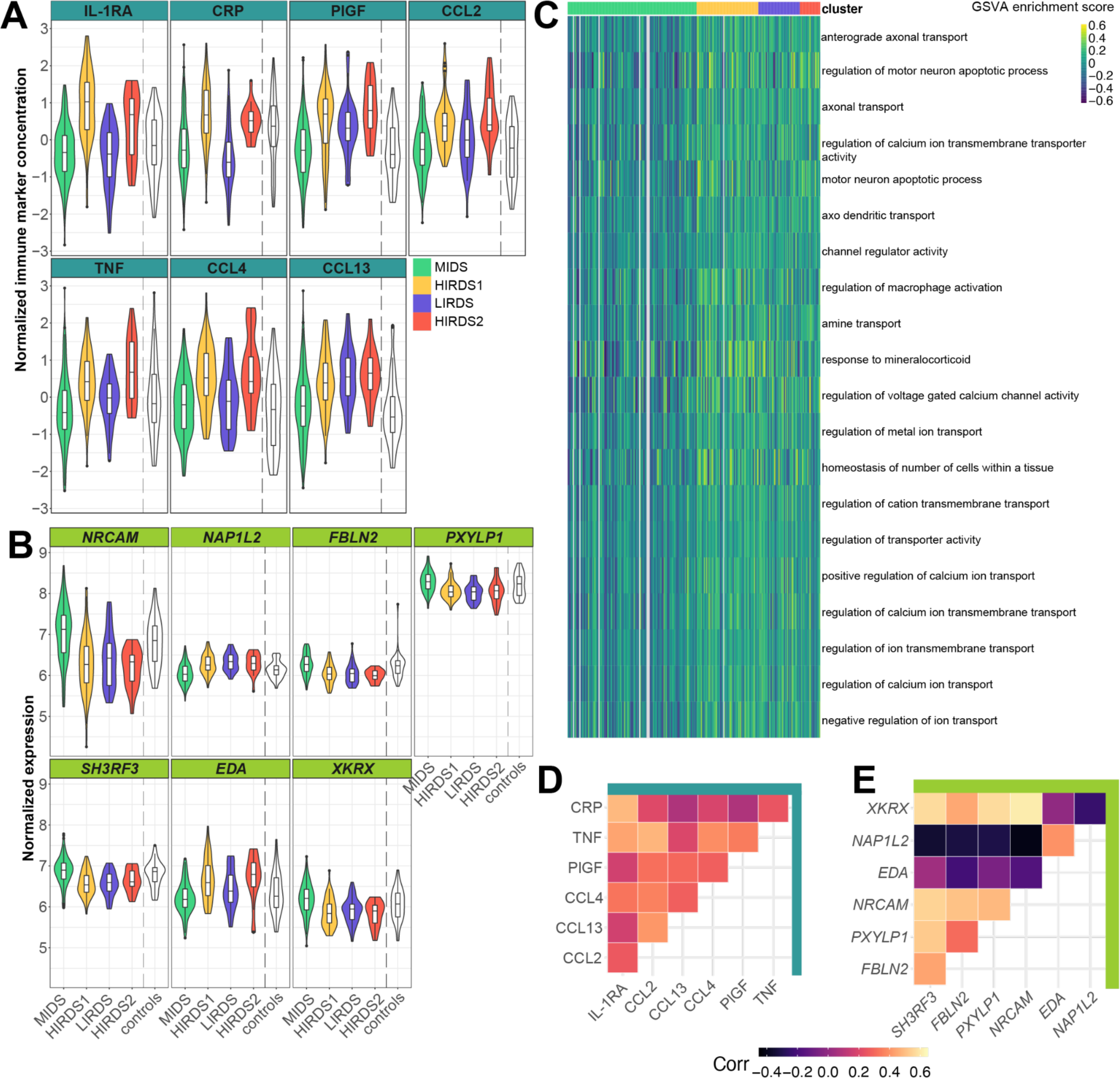
Immune marker and RNA-seq data contains complementary information in the initial clustering with age, BMI, immune markers and RNA-seq data. (A) Violin plots showing the distribution of the seven most differentiating immune markers across the clusters. (B) Violin plots showing the distribution of the seven most differentiating genes across the clusters. (C) Heatmap of the 20 most differentiating GO gene sets ordered by their variable importance that are enriched in the clusters. These gene sets were identified through Gene Set Variation Analysis (GSVA). (D) Heatmap showing the Pearson correlation between the top immune markers. (E) Heatmap showing the Pearson correlation between the top genes. MIDS = mild depression symptoms cluster, HIRDS = high immune-related depression symptoms cluster, LIRDS = low immune-related depression symptoms cluster.

In contrast, the most discriminating genes mainly differentiated the MIDS cluster, defined by a low depression severity, from the other clusters (Figure 4B). Accordingly, the correlations between the genes were stronger than those among immune markers, with the exception of the correlation between *EDA* and *NAP1L2*. On average, the absolute correlation between the top genes was 0.36, while it was 0.27 between the top immune markers (Figure 4E). To link the findings to clinical data, we investigated if the top discriminating genes could predict the responder status in patients with depression to treatment with an antibody against TNF in an external data set (Mehta et al. 2013). Based on the area under the curve (AUC), the top 10 and top 20 genes did not perform better than random genes (permutation test, p = 0.139 and p = 0.159). The most important gene for the 20-gene-model was vesicle associated membrane protein 1 (*VAMP1*), which was also included in the 10-gene prediction model, although it was not statistically significant.

To understand the underlying molecular mechanisms, we analyzed not only individual genes but also their coordinated expression patterns, examining these genes in the context of pathways. We applied gene set enrichment analysis using GSVA (Hänzelmann, Castelo, and Guinney 2013) to calculate gene set scores for every participant and compared the distribution across clusters (Table S8). As depicted in Figure 4C, the gene sets that stood out prominently were enriched in the LIRDS and HIRDS clusters. Interestingly, these gene sets included those related to the brain (e.g., anterograde axonal transport, VI = 17.3, 38 genes), immune system (regulation of macrophage activation, VI = 12.5, 32 genes), stress response (response to mineralocorticoid, VI = 12.4, 15 genes) and (ion) transport (e.g., regulation of calcium ion transmembrane transporter activity, VI = 13.1, 44 genes).

### 3.2 Re-evaluating the initial clustering adjusted for age, BMI and sex showed no phenotypic differences

Considering the strong impact of BMI and age on the clustering outcomes, we re-evaluated the clustering by first adjusting the biological data for age, BMI and sex using a linear model, and then applying the clustering algorithm. After adjusting the data, we obtained a three cluster solution (Figure S2C-D), which differed from the four clusters identified in the initial analysis (Figure S8A). Notably, in the re-evaluated clustering, the variable importance of the immune markers was diminished relative to the RNA-seq data (Table S9). Among the immune makers, IL-27 stood out with the highest immune marker variable importance of 7.4. Contrastingly, the gene CKLF like MARVEL transmembrane domain containing 6 (*CMTM6*) showed a much higher variable importance of 77.4. Moreover, the primary discriminative immune markers or genes in this analysis were different from those in our initial clustering (Figure S8B). Compared to the genes, the distribution of the immune markers across the clusters was more uniform (Figure S8C-D). There were no significant differences of age, BMI, BDI or MADRS sum scores between the clusters (Figure S8E-G).

### 3.3 Re-evaluating the initial clustering including cell types identified three clusters predominantly driven by RNA-seq data

Given the previous findings that highlight the significance of blood cell type composition in discerning depression subgroups (Lynall et al. 2020), we integrated cell types into a new clustering model aligning them in one layer with age and BMI. We did not have cell counts available but deconvoluted cell type proportions from the RNA-seq data. The predominant cell types in our sample were “T cells CD4 memory resting” (median fraction of 0.41) and “Macrophages M1” (median fraction of 0.15, see Figure 5A). The new clustering led to a three cluster solution (Figure S2E-F). Cluster 1 contained mostly (85.7%) individuals that were assigned to the MIDS cluster in the initial clustering without the inclusion of cell types. Cluster 3 contained predominantly individuals from the clusters with high BMI and high BDI (HIRDS clusters, 69.2%) as determined from the initial clustering. Cluster 2 presented a mix, drawing from different groups of the initial clustering: 54.2% from MIDS, 17.7% from HIRDS1, 24% from LIRDS and 4.2% from HIRDS2 (Figure 5B). Accordingly, we termed the clusters reMIDS, *intermediate depression symptoms* cluster (reIDS) and reHIRDS.

**Figure 5:**
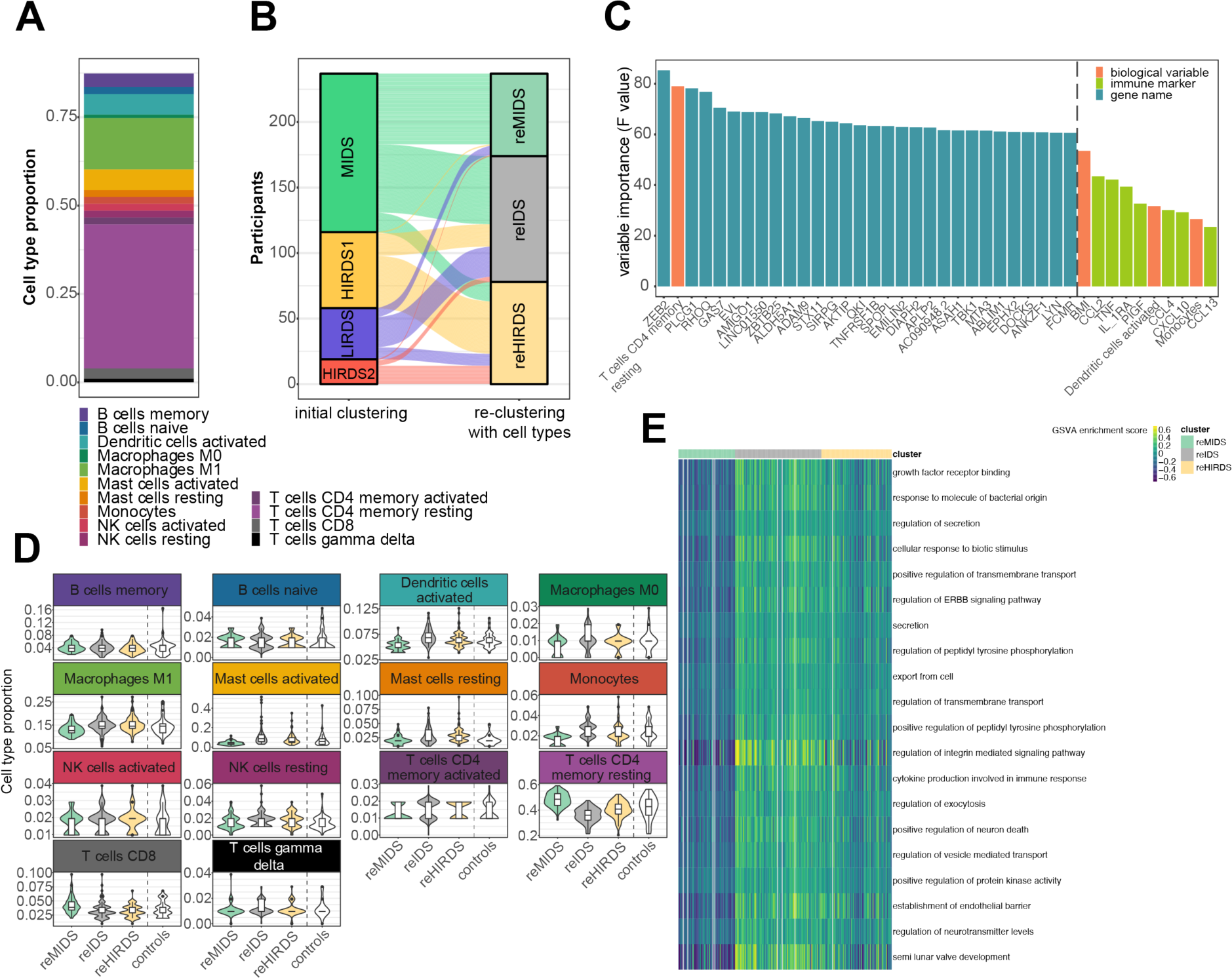
Re-clustering with age, BMI, cell types, immune markers and RNA-seq data. (A) Stacked barplot illustrating the proportion of various cell types predicted from the RNA-seq data for all participants. (B) Mapping of the participants from the initial clustering to the re-evaluated clustering that additionally contains cell type proportions. (C) Barplot showing the variable importance of the top 30 variables that most differentiate the clusters, along with the 10 most differentiating biological variables and immune markers. (D) Violin plots illustrating the predicted cell type proportions across the clusters. (E) Heatmap of the 20 most differentiating GO gene sets, ordered by their variable importance, that are enriched in the clusters. These gene sets were identified through Gene Set Variation Analysis (GSVA). reMIDS = re mild depression symptoms cluster, reIDS = re intermediate depression symptoms cluster, reHIRDS = re high immune-related depression symptoms cluster, NK = natural killer cells.

#### 3.3.1 T cell proportions and RNA-seq data were important for the clustering

The distribution of age and BMI demonstrated an increasing trend across the clusters, especially BMI was markedly increased in the reHIRDS cluster, with 29.1 compared to 22 for the reMIDS cluster and 23.7 for the reIDS cluster (Figure S9A-B). In line with the pattern seen in the initial clustering, the reHIRDS cluster showed significantly elevated BDI compared to the other clusters (Tukey HSD test with adjusted p values, p = 0.000126 for cluster 1 and p = 0.008659 for cluster 2, Figure S9C), and the MADRS in the reHIRDS cluster was elevated compared to the reMIDS cluster (Tukey HSD test with adjusted p values, p = 0.013896) as well.

In reassessing the variables contributing to the clustering, we observed a shift from our initial findings. While age and BMI were the most important variables in the initial clustering, their impact reduced in the re-evaluation. BMI still remained the second most important biological variable (VI = 53.5, Figure 5C and Table S10), ranking 77th overall, but age (VI = 19.5) dropped to the 2037th position. The top 7 immune markers were the same as in the initial clustering except CXCL10 (IP-10, VI = 29.3) replacing CRP (VI = 21.3). However, in the reclustering the immune markers ranked 240th to 1484th in the variable importance, whereas in the initial clustering 3rd to 24th. The cell type “T cells CD4 memory resting” emerged as the most important biological variable (VI = 79.1), also being the most prevalent cell type in our sample (Figure 5A). Other notable cell types included “Dendritic cells activated” (VI = 31.7) and “Monocytes” (VI = 26.6). In the reMIDS cluster, “T cells CD4 memory resting” showed an increased mean proportion of 0.48 compared to 0.36 and 0.40 in the other clusters, while the “Dendritic cells activated” and “Monocytes” were decreased in the reMIDS cluster (Figure 5D). Furthermore, Figure 5C shows that genes were the essential variables to discriminate between the clusters (see Supplementary Results for more details).

#### 3.3.2 Pathway analysis identified immune system involvement

Next, we performed gene set enrichment analysis to analyze the biological pathways that differed between the cluster. As highlighted in Figure 5E, the most discriminative gene sets were downregulated in the reMIDS cluster, upregulated in the reIDS cluster and moderately upregulated in the reHIRDS cluster. Closer examination revealed that these gene sets were enriched for terms related to the immune system (response to molecule of bacterial origin, VI = 119, 186 genes), secretion (regulation of secretion, VI = 118.1, 297 genes), transmembrane transport (regulation of transmembrane transport, VI = 108, 237 genes) and neurons (positive regulation of neuron death, VI = 103.7, 58 genes) among others (Table S11).

### 3.4 Integration of protein and RNA-seq data with single cell expression showed cell type specific regulation of inflammation

To further elucidate the characteristics of the high depression severity clusters from our initial clustering model delineated in Figure 2, we analyzed the genes and immune markers that differentiated the LIRDS and HIRDS clusters. Two prominent variables emerged: the gene serpin family F member 1 (*SERPINF1*, VI = 9.1) and the protein vascular endothelial growth factor A (VEGF-A, VI = 10.1). As represented in Figure 6A and 6B, *SERPINF1* expression was elevated in the MIDS and LIRDS clusters, coinciding with low VEGF-A protein concentrations. In contrast, the HIRDS clusters displayed an opposite pattern. Interestingly, we did not find differences in the gene expression of *VEGFA* between the clusters (Figure 6C) and in general low correlation between RNA-seq and corresponding protein level (see Supplementary Results and Figure S10 for more details).

**Figure 6:**
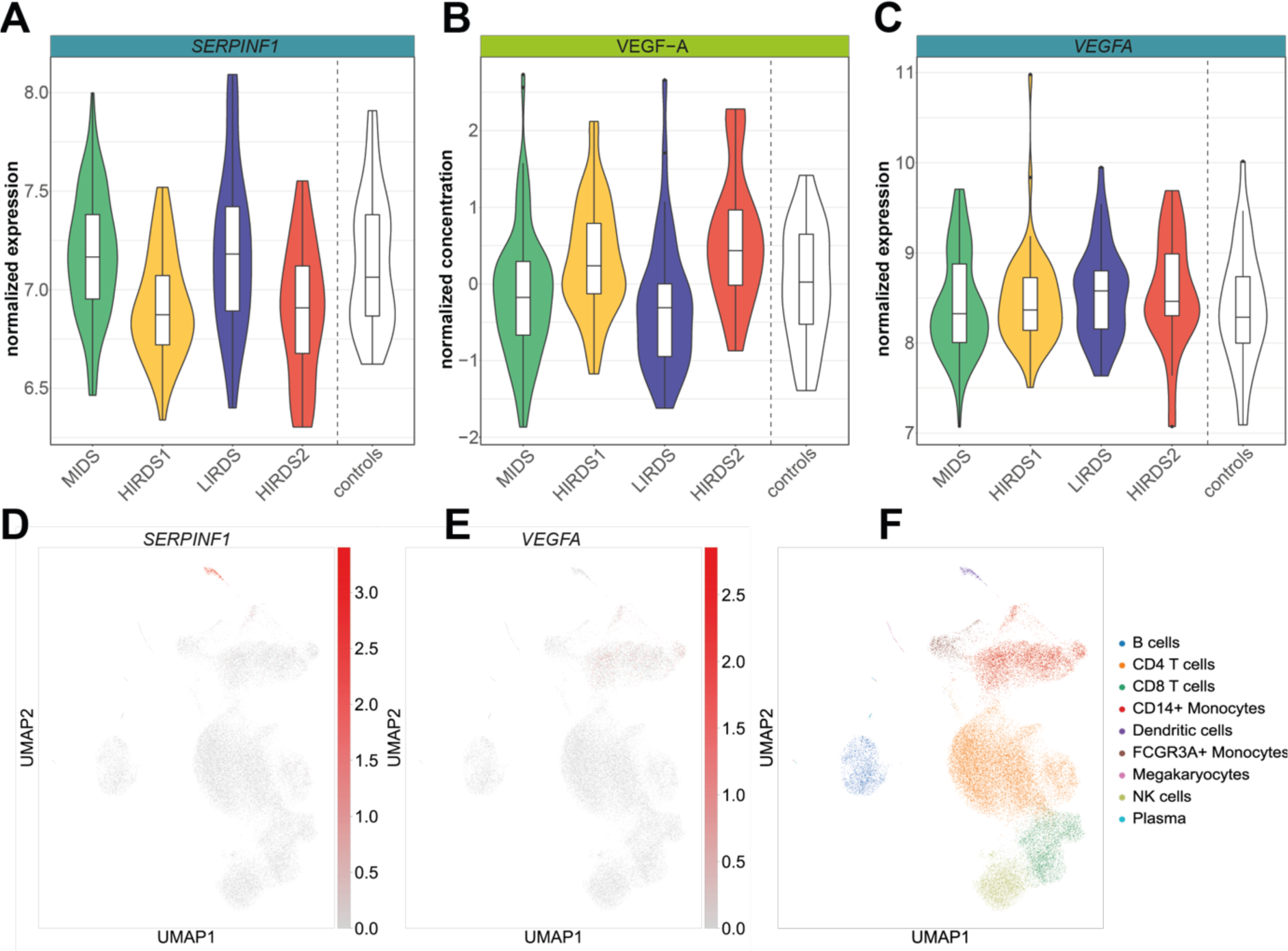
Cell-type specific dysregulation in the initial clustering with age, BMI, immune markers and RNA-seq data. Violin plots displaying: (A) the gene expression of SERPINF1 across the clusters, (B) normalized protein concentration of VEGF-A across the clusters, and (C) gene expression of VEGFA across the clusters. (D) - (F) Single-cell RNA-seq data of peripheral blood mononuclear cells (PBMCs) from 14 patients with depression and other mental disorders visualized using UMAP plots. (D) Demonstrates the gene expression of SERPINF1. (E) Shows the gene expression of VEGFA. (F) Illustrates the cell types identified within the PBMCs. NK = natural killer cells, MIDS = mild depression symptoms cluster, HIRDS = high immune-related depression symptoms cluster, LIRDS = low immune-related depression symptoms cluster.

The importance of different cell types was underlined by single cell gene expression analysis. PBMC RNA-seq data from 14 individuals with depression and other mental disorders indicated a predominant *SERPINF1* expression in dendritic cells (Figure 6D and 6F), while *VEGFA* was mainly expressed in monocytes (Figure 6E-F), showing the importance of cell types even when not included directly in the clustering.

## 4. Discussion

In this study, we analyzed a transdiagnostic sample of individuals with stress-related mental disorders from two cohorts to identify patterns of depression symptoms in relation to immune alterations, thereby broadening the understanding of immune-related subgroups in depression. Traditionally, such subgroups have been defined by specific immune markers such as CRP or TNF, known to be increased in some depressed patients (Haapakoski et al. 2015; Osimo et al. 2020), especially those resistant to antidepressant treatment (Chamberlain et al. 2019; Yang et al. 2019; Strawbridge et al. 2015). Our large-scale multi-omics analysis, integrating gene expression data of 12210 genes with immune marker profiling of 43 makers including CRP, suggests a more complex immune involvement in depression symptoms than indicated by CRP alone. Our approach uncovered four distinct clusters dissecting depression symptoms across the diagnostic spectrum: two *high immune-related-depression symptoms* (HIRDS) clusters with increased immune markers, BMI, and depression severity; a *mild depression symptoms* (MIDS) cluster, predominantly younger participants with lower depression severity and minimal immune marker elevation; and a *low immune-related-depression symptoms* (LIRDS) cluster with elevated depression severity but without increased immune markers. This nuanced classification offers deeper insights into the interplay between immune function and depression symptoms.

Incorporating a multi-omics integration clustering approach has clearly demonstrated its value, providing unique insights through the combination of phenotypic data, immune markers and RNA-seq data. This was especially evident in identifying HIRDS clusters with elevated immune markers such as CRP, TNF, IL-1RA, PlGF, CCL2, CCL4, and CCL13.

Measuring IL-1 beta posed a challenge due to its low concentration, even with high-sensitivity assays, highlighting the difficulties in detecting certain cytokines. Interestingly, IL-1RA, typically anti-inflammatory, is produced under the same inflammatory conditions as the proinflammatory IL-1 beta, linking it to an increased risk of developing depressive symptoms in elderly (Osimo et al. 2020; Milaneschi et al. 2009). Consistent with this, we found heightened IL-1RA levels in our HIRDS clusters, which also exhibit altered appetite patterns, aligning with findings from Simmons et al. 2020. While specific BDI items like increased appetite and hypersomnia showed only a trend, a clear elevation in overall depression severity and BMI was observed. These patterns support the concept of immuno-metabolic depression, where heightened inflammation correlates with disrupted energy balance, manifesting as obesity and fatigue (Lamers et al. 2020; Milaneschi et al. 2020; Simmons et al. 2020).

Our research sheds light on the role of less-studied chemokines in depression. Elevated CCL2 in blood, CSF, and in post-mortem brain tissues of depressed patients (Young, Bruno, and Pomara 2014; Eyre et al. 2016; Sørensen et al. 2023; Torres-Platas et al. 2014), alongside mixed data for CCL4 (Camacho-Arroyo et al. 2021; Sørensen et al. 2023; Leighton et al. 2018) and limited research on CCL13, with only one study that found lower levels in suicide attempters (Janelidze et al. 2013), indicate their intricate roles in this symptom spectrum. Additionally, it highlighted PlGF, part of the vascular endothelial growth factor family (De Falco 2012), implicated in angiogenesis, the immune response, and obesity-related processes (Oura et al. 2003; Lijnen et al. 2006), which was previously found to have lower levels in depressed patients (Yue et al. 2016). By employing a multi-panel approach to simultaneously measure a wide array of immune markers, we elucidated the complex roles of chemokines and established markers in depression symptom spectrum. This strategy not only identifies specific immune markers associated with depression symptoms but also advances our understanding of the disorder’s immunological aspects, suggesting a promising direction for future research.

The immune markers were crucial for differentiating between the HIRDS clusters and others, while key genes and the gene set analysis were instrumental in separating between clusters of varying depression severity. The panel of important genes includes genes that are associated with neurodevelopmental disorders (*NRCAM*, Kurolap et al. 2022), neuronal development (*NAP1L2*, Attia et al. 2007) or that were downregulated in post mortem choroid plexus from patients with MDD (*FBLN2*, Turner et al. 2014). This underlines that depression not only affects the brain but other tissues as well and demonstrates our approach’s effectiveness at identifying genes differentiating depression severity. Our gene set analysis further supports this, revealing a prevalence of pathways related to neurons and axonal transport in the high depression severity clusters. While immune system pathways were enriched in the genes differentiating the clusters, the immune marker data primarily defined the immune-related depression symptom clusters, underscoring the value of this integrative approach.

The significance of BMI and age in relation to depression symptoms in our analysis added to the ongoing debate about whether BMI adjustments in immune marker analyses clarify or confound the relationship with depression (Horn et al. 2018; Moriarity, Mengelkoch, and Slavich 2023). Several studies suggest that CRP’s association with depression is more pronounced without BMI-adjustments (Horn et al. 2018; Chae et al. 2022; Figueroa-Hall et al. 2022). Recently, a simulation study demonstrated that including BMI as a covariate can lead to reduced precision in estimating the relationship of inflammation on depression (Moriarity, Mengelkoch, and Slavich 2023). Consistently, our findings revealed no significant differences in depression severity or inflammation status when adjusting for BMI, sex and age, underscoring the complex interplay among these factors and emphasizing BMI’s role as an integral component of the depression phenotype.

Our analysis extended beyond clinical depression severity to heart rate variability and neuroimaging, differentiating the clusters. While not corrected for covariates, heart rate variability (SDNN) was lower in the LIRDS and HIRDS clusters and could discriminate them from the MIDS cluster, aligning with previous findings that link reduced heart rate variability to current and former depression, as well as dysphoria (Hartmann et al. 2019; Koch et al. 2019; Dell’Acqua et al. 2020), which suggests broader physiological implications of depression. Structural neuroimaging revealed a trend of relatively higher total brain volume in the LIRDS cluster compared with the HIRDS and MIDS clusters, with similar effects for GM and WM analyzed separately. Regional analyses confirmed inverse changes of cerebrospinal fluid (CSF), specifically of the lateral ventricle volume. In the presence of acute depressive symptoms, it seems that the LIRDS constellation might protect from CSF including ventricular enlargement. Larger ventricles have been observed in MDD with early disease onset before 21 years of age (Schmaal et al. 2016) and in bipolar disease (Hibar et al. 2016) which could suggest that stronger immune activation might take influence on subclinical or masked bipolar traits. Regional effects as detected for the bilateral hippocampi and the right rostral anterior cingulate cortex, were heterogeneous between the two HIRDS clusters. While these structures were reported before in immune based stratifications (Savitz et al. 2013) and are plausibly involved in stress response regulation (Herman et al. 2016), larger samples are needed to fully map the anatomical characteristics of immune-related depression.

Our analysis supports the hypothesis that depression is associated with alterations in cell type composition and immune marker levels (Lynall et al. 2020; Foley et al. 2022). Integrating cell type proportions refined our initial clusters, revealing three re-clusters with varying depression severity and aligning well with the initial findings. This highlights the robustness of our subgroups. The reHIRDS cluster, marked by elevated BMI and immune markers, showed an intermediate alteration in T cells CD4 memory resting proportion deviating from Foley et al. 2022 who reported no significant CD4+ T cells proportion changes in depression. Conversely, our reHIRDS and *intermediate depression symptoms* (reIDS) clusters reflected their findings of decreased lymphocyte percentage in depression. In contrast to the results from Lynall et al. 2020 who observed elevated immune markers and absolute immune cell counts in their immune-related depression cluster, our reHIRDS cluster did not exhibit the strongest cell type phenotype. This discrepancy could stem from methodological differences, such as direct cell counts versus deconvolution. This suggests a complex interplay between immune markers and PBMC composition in depression. Possible sources of immune markers like neutrophils (Tecchio, Micheletti, and Cassatella 2014) and adipose tissue (Fain 2006; Shelton et al. 2015; Shelton and Miller 2011) highlight the importance of obesity in immune-related depression. Our findings emphasize the need for further research into the associations of immune cell composition and immune markers in this context.

The advantages of a multi-omics integration were further demonstrated by revealing cell type specific inflammation of immune-related depression symptom clusters. VEGF-A was elevated in the HIRDS clusters, contrasting the *SERPINF1* expression, which showed the opposite pattern. This aligns with its gene product, pigment epithelium-derived factor (PEDF), known to inhibit VEGF-A (Dawson et al. 1999). Of note, PEDF is neurotrophic (Tombran-Tink and Barnstable 2003) and was shown to ameliorate depression-like symptoms in mice (Tian et al. 2020) and was increased in depression patients after electroconvulsive treatment (Ryan et al. 2017). This suggests a link of the dysregulated immune pathway in the HIRDS clusters and the observed elevated depression severity.

Furthermore, our single cell data revealed that *SERPINF1* is mainly expressed in dendritic cells, suggesting a potential reduction of these cells within immune-related depression symptom clusters. This finding merits additional exploration, as it could open new pathways for understanding and potentially targeting the immune-related aspects of depression symptoms.

Our study has some limitations worth noting. Firstly, the heterogeneity of our sample, which includes a transdiagnostic sample of both in- and out-patients with varying medication usage and comorbid conditions, may affect the generalizability of our findings. Secondly, the imaging and psychophysiology data represents a proportion of the total samples (63%), which requires caution in interpretation of these findings. Additionally, the cell type composition in our study was inferred from the RNA-seq data rather than being directly measured, which could impact the accuracy of these estimates. As there was no validation sample, these limitations underscore the importance of further research to validate and expand our findings.

Taken together, our multi-omics integration analysis successfully discovered two clusters with immune-related depression symptoms, supporting the immuno-metabolic depression hypothesis and highlighting the importance of biological variables such as age and BMI. Incorporating single cell data, we uncovered cell type specific inflammation dysregulation involving *SERPINF1* and VEGF-A, both previously implicated in depression. This integrated approach, recognizing depression’s heterogeneity, enhances our understanding of its complexity by exploring symptoms across diagnoses. It highlights novel immune markers and genes as potential targets for clinical stratification and new therapeutic intervention.

## Data and Code availability

The computational code has been made available on GitHub: https://github.com/cellmapslab/ImmuneDepressionSymptoms, while the RNA-seq data is accessible in the GEO repository under GSE260603 and the immune marker data in the Zenodo repository https://zenodo.org/doi/10.5281/zenodo.10698978.

## Conflict of interest

VIS has provided consulting and advisory services for Roche and Sony. The other authors declare no conflict of interest.

## Data Availability

https://www.ncbi.nlm.nih.gov/geo/query/acc.cgi?acc=GSE260603

## Acknowledgments

We extend our gratitude to all participants of the BeCOME and OPTIMA studies without whom our research would not be possible. We thank the Biomaterial processing and repository unit (BioPrep) of the Max Planck Institute of Psychiatry for their handling of the biological samples and the RNA isolation.

Dr. Knauer-Arloth’s contributions were supported by the Brain & Behavior Research Foundation (NARSAD Young Investigator Grant, #28063).

## Contributions

Jonas Hagenberg: Conceptualization, Methodology, Software, Formal analysis, Writing - Original Draft, Visualization

BeCOME study group: Resources OPTIMA study group: Resources

Tanja Brückl: Data curation, Formal analysis, Writing - Review & Editing Mira Erhart: Formal analysis, Writing - Review & Editing

Johannes Kopf-Beck: Data curation, Writing - Review & Editing Maik Ködel: Investigation

Ghalia Rehawi: Software, Writing - Review & Editing Simone Röh-Karamihalev: Software

Susann Sauer: Investigation

Natan Yusupov: Data curation, Writing - Review & Editing Monika Rex-Haffner: Investigation

Victor Spoormaker: Supervision, Writing - Review & Editing

Philipp Sämann: Methodology, Formal Analysis, Writing - Review & Editing Elisabeth Binder: Supervision, Funding acquisition, Writing - Review & Editing

Janine Knauer-Arloth: Conceptualization, Visualization, Supervision, Funding acquisition, Writing - Review & Editing

JH processed the immune marker and RNA-seq data, designed and carried out the clustering and the subsequent analyses, provided critical intellectual input and wrote the initial manuscript draft; BSG and OSG recruited participants, carried out the studies and provided data; TMB processed the phenotype data and provided critical intellectual input on the phenotypes; ME and VIS led the electrocardiography data analysis; JKB provided phenotype data; MK performed immune marker measurements; GR advised on cell type deconvolution; SRK advised on RNA-seq data analysis; NY processed phenotype data; MRH supported experimental procedures and project organization; PS led the imaging data analysis and contributed to the manuscript draft; EB supervised the project and acquired funding; JKA conceived the idea, acquired funding, supervised the project, designed experiments and analyses, provided critical intellectual input and contributed to the manuscript draft writing.

## Notes

### Author Declarations

Ethics committee of Ludwig-Maximilians-University Munich, Germany gave ethical approval for this work

