## Supplemental material for "Dissecting depression symptoms: multi-omics clustering uncovers immune-related subgroups and cell-type specific dysregulation"

### 1. Supplementary Methods

#### 1.1 Sample selection

The study sample comprised 246 participants from the Biological Classification of Mental Disorders (BeCOME) study (ClinicalTrials.gov: NCT03984084, Bruckl et al. 2020) and 115 participants from the OPtimized Treatment Identification at the MAx Planck Institute (OPTIMA) study (ClinicalTrials.gov: NCT03287362, Kopf-Beck et al. 2020). The BeCOME study contains participants with and without mental disorders, mostly outpatients and participants with no psychopharmacological treatment for a minimum of 2 months before entering the study, while the OPTIMA study contains inpatient and day clinic patients admitted for the treatment of a major depressive episode who are mostly under psychopharmacological treatment. The Munich-Composite International Diagnostic Interview (DIA-X/M-CIDI) (Wittchen et al. 1995; Wittchen and Pfister 1997) was employed to assess all participants. The DIA-X/M-CIDI is a modified version of the World Health Organization CIDI, version 1.2 supplemented by questions to cover DSM-IV and ICD-10 criteria. Out of the participants with immune marker measurement, 237 affected participants (BeCOME=134, OPTIMA=103, Figure S1) met either threshold or subthreshold (falling short of one DSM-IV criterion such as the impairment criterion) DSM-IV DIA-X/M-CIDI criteria for any substance use, affective or anxiety disorder including post-traumatic stress disorder and obsessive-compulsive disorder within the last 12 months. 192 of these participants had a DSM-IV diagnosis of major depression or dysthymia. We also included 36 mentally healthy participants from the BeCOME study without any DSM-IV IDA-X/M-CIDI diagnosis as controls.

#### 1.2 Ethics approval and informed consent

The BeCOME and OPTIMA studies were approved by the ethics committee of the Ludwig Maximilian University, in Munich, Germany, under the reference numbers 350–14 and 17–395, respectively. Written informed consent was obtained from all participants before study enrollment.

#### 1.3 Assessments

##### 1.3.1 Questionnaire data

All participants were assessed by the Beck Depression Inventory (BDI) II (Hautzinger, Keller, and Kühner 2006) and the Montgomery–Åsberg Depression Rating Scale (MADRS, Schmidtke et al. 1988). An overview about the sample characteristics for this subset are provided in Table 1, for more details see Table S1.

##### 1.3.2 Blood collection

Blood for plasma was collected in 7.5ml EDTA-plasma tubes (Sarstedt, Nümbrecht, Germany, Cat. No. 01.1605.001) in the morning under fasted conditions and centrifuged at 2500 rcf at 4°C for 15 minutes (Model 5810 R, Eppendorf, Hamburg, Germany). Plasma was aliquoted in 0.5ml Cryo Tubes (Azenta, Burlington, USA, Cat. No. 68-0703-12) and stored at -80°C.

Blood for peripheral blood mononuclear cells (PBMCs) was collected in 3 x 9ml EDTA tubes (Sarstedt, Nümbrecht, USA, Cat. No. 02.1066.001). PBMCs were extracted from diluted whole blood using Biocoll (Bio&SELL, Feucht, Germany, Cat. No. BS.L.6115) for a gradient centrifugation in 50ml SepMate Tubes (Stemcell Technologies, Vancouver, Canada, Cat. No. 85460). Density gradient centrifugation was performed with the brake on, after which isolated PBMCs were transferred into a fresh tube. The cell pellet was washed twice and the number of cells was determined with the TC20 Automated Cell Counter (BioRad, Hercules, USA).

After renewed centrifugation and aspiration of the supernatant, cells were resuspended in a freezing medium (50 % FBS, 40 % RPMI, 10 % DMSO). 10Mio cells per ml were aliquoted in 1.9ml Cryo Tubes (Azenta, Burlington, USA, Cat. No. 65-7643) and stored in the gas phase above liquid nitrogen.

##### 1.3.3 Immune marker measurements

We used the V-PLEX Human Biomarker 54-Plex Kit (Meso Scale Diagnostics (MSD), Rockville, USA, Cat. No. K15248G-2) to measure immune markers in plasma. The samples were randomized into 96 well plates. The plates were analyzed with the MESO QuickPlex SQ 120 imager. In addition, enzyme-linked immunosorbent assay (ELISA) was used to measure the following markers: high-sensitivity C-reactive protein (hsCRP, Tecan Group Ltd., Männedorf, Switzerland, Cat. No. EU59151), cortisol (Tecan Group Ltd., Männedorf, Switzerland, Cat No. RE52061), interleukin (IL)-6 (Thermo Fisher Scientific, Waltham, USA, Cat. No. BMS213HS), IL-6 soluble receptor (sIL-6R, Thermo Fisher Scientific, Waltham, USA, Cat. No. BMS214) and IL-13 (Thermo Fisher Scientific, Waltham, USA, Cat. No. BMS231-3). All assays were performed according to the manufacturer’s instructions. Immune markers with more than 16% missing values or a high sensitivity alternative were excluded (p = 17 markers). Values below the detection limit in markers measured with ELISA were set to zero and values above the detection limit to the upper limit. 15 participants with a hsCRP concentration higher than 20 mg/L were excluded from the sample as this indicates an acute infection. The remaining 43 markers were: fibroblast growth factor 2 (FGF2 or bFGF), cortisol, C-C motif chemokine 11 (CCL11 or eotaxin), CCL26 (eotaxin-3), vascular endothelial growth factor receptor 1 (VEGFR1 or Flt-1), hsCRP, intercellular adhesion molecule (ICAM)-1, interferon (IFN)-gamma, IL-1alpha, IL-1 receptor antagonist (IL-1RA), IL-10, IL-12/IL-23p40, IL-12p70, IL-13, IL-15, IL-16, IL-17A, IL-17B, IL-2, IL-27, IL-31, IL-5, IL-6 high sensitivity (IL-6HS), IL-7, IL-8HS, C-X-C motif chemokine 10 (CXCL10 or IP-10), CCL2 (MCP-1), CCL13 (MCP-4), CCL22 (MDC), CCL3 (MIP-1alpha), CCL4 (MIP-1beta), placental growth factor (PlGF), serum amyloid A (SAA), sIL-6R, CCL17 (TARC), angiopoietin-1 receptor (Tie-2), tumor necrosis factor (TNF or TNF-alpha), lymphotoxin-alpha (LT-alpha or TNF-beta), thymic stromal lymphopoietin (TSLP), vascular cell adhesion protein 1 (VCAM-1), vascular endothelial growth factor (VEGF)-A-HS, VEGF-C, VEGF-D. For an overview about missing values, see Table S2.

##### 1.3.4 RNA extraction

The frozen samples were randomized. After thawing at 37°C, 200µl of the PBMCs were dissolved in 5ml PBS, centrifuged, supernatant removed and again centrifuged with 5ml PBS. The supernatant was removed, the sample resuspended in 300µl water, added to 300µl lysis buffer in deepwell plates and stored at -80°C until further processing. The RNA was extracted automatically with the chemagic 360 instrument (PerkinElmer, Waltham, USA) by using the chemagic RNA Kit (PerkinElmer, Waltham, USA, Cat. No. CMG-1212) according to the manufacturer’s instructions and 25µl eluate stored in a PCR plate at -80°C until further processing. The quality of the RNA was assessed with the 2200 TapeStation (Agilent, Santa Clara, USA).

##### 1.3.5 RNA-sequencing

200ng RNA was used for rRNA depletion with the RiboCop rRNA Depletion Kit V1.3 (Lexogen, Vienna, Austria, Cat. No. 037.96). 10µl of the depleted RNA was used for library preparation with the Lexogen CORALL total RNA-Seq Library Prep Kit with UDIs 12 nt Sets A1-A4 (Cat. No. 117.96, 132.96, 133.96, 134.96). A qPCR with the Lexogen PCR Add-on Kit for Illumina (Cat. No. 020.96) was performed to determine the optimal PCR cycle number for the endpoint PCR. The DNA content of the libraries were measured with the Qubit fluorometer Q32857 (Invitrogen, Carlsbad, USA) and all libraries were pooled equimolar into one library. Afterwards, a free adapter blocking treatment was performed with Illumina Free Adapter Blocking Reagent Kit (Cat. No. 20024145). The quality of the final library was assessed with the 2100 Bioanalyzer (Agilent, Santa Clara, USA) and the KAPA Library Quantification Kit (Roche Diagnostics, Rotkreuz, Switzerland, Cat. No. 07960298001) on the Roche LightCycler 480. A total of 160µl from the prepared libraries, each with a concentration of at least 5nM, were sequenced on an NovaSeq 6000 (Illumina, San Diego, USA). The sequencing was conducted using an S4 200 flow cell with paired-end 100bp reads, yielding an average of 30.6 million paired reads per library.

##### 1.3.6 Structural magnetic resonance imaging (MRI) data assessment

Structural MRI analysis was performed on 151 affected participants (BeCOME=117 and OPTIMA=34) and 29 healthy controls after exclusion of 7 participants based on radio-pathological findings. High resolution T1-weighted images that had the identical sequence in both original studies (Bruckl et al. 2020; Kopf-Beck et al. 2020) (Sagittal FSPGR 3D BRAVO, TE 2.3 ms, TR 6.2 ms, TI 450 ms, FA 12°, FOV 25.6 × 25.6 × 20.0 cm3, matrix 256 × 256, voxel size 1 × 1 × 1 mm3) underwent two processing streams: (1) Automated cortical and subcortical segmentation using FreeSurfer v7.1.1 (Fischl 2012), after a pre-step to attenuate the MR coiled-based bias field using SPM12. Quality control (QC) was based on visual steps following standard procedures developed in the ENIGMA consortium (<https://enigma.ini.usc.edu/protocols/imaging-protocols>). None of the 151 participants showed signs of fault segmentations. (2) Voxel-based segmentation using CAT12 (<https://neuro-jena.github.io/cat>) to gain MNI normalized and Jacobian modulated gray matter (GM), white matter (WM) and cerebrospinal fluid (CSF) maps (isometric voxels 1.5 mm). QC comprised visual inspection of all GM maps and verifying image quality ratings ≥ 0.7 (Gaser et al. 2022). Eventually, GM maps were spatially smoothed (Gaussian kernel, FWHM 6 × 6 × 6 mm3) for voxel-based morphometry (VBM).

##### 1.3.7 Electrocardiography data assessment

A subset of the participants had an electrocardiography (ECG) recording that spanned 24h and was conducted at the first day of the BeCOME and within the first week (baseline assessment before treatment) of the OPTIMA study. The recordings were done with the portable ECG-device Faros 180 (Mega Electronics Ltd, Kuopio, Finland) at a sampling frequency of 500 Hz. After exclusions due to the absence of a sleeping period within the 24h recordings, a subset of 149 affected participants (BeCOME=180 and OPTIMA=41) were included in the subset and another 32 healthy controls.

#### 1.4 Data analysis

##### 1.4.1 Immune marker analysis

The immune markers were quantile-normalized, wherein values were ranked and then mapped to the quantiles of a standard normal distribution. Batch variables were assessed by ANOVA for every immune marker and the data was corrected for the biobank storage position. For each immune marker, a linear model was fitted in R version 4.0.2 (R Core Team 2023) to regress out the batch variable. The residuals from this regression were then used for downstream analysis.

##### 1.4.2 RNA-seq analysis

The RNA-seq data quality was assessed with FastQC version 0.11.8 (Andrew 2010) and the Unique Molecular Identifiers extracted with UMI-tools version 1.1.1 (Smith, Heger, and Sudbery 2017). Next, cutadapt version 2.10 (Martin 2011) was used to trim the adapters. The sequences AGATCGGAAGAGCACACGTCTGAACTCCAGTCA and AGATCGGAAGAGCGTCGTGTAGGGAAAGAGTGT were used for read 1 and read 2, respectively. The reads were aligned to GRCh38.p12 (Ensembl version 97, Martin et al. 2023) with STAR version 2.7.7a (Dobin et al. 2013). Afterwards, samtools version 0.1.19 (Danecek et al. 2021) was used to create indices of the bam files and UMI-tools version 1.1.1 to deduplicate reads. Picard version 2.20.4 (Broad Institute 2019) was used to calculate RNA metrics and mark duplicates. Finally, the mapped reads were counted with featureCounts version 1.6.4 (Liao, Smyth, and Shi 2014) and the metrics summarized with MultiQC version 1.9 (Ewels et al. 2016).

RNA-seq data was available for 356 participants. We excluded nine participants due to a mismatch between the RNA-seq data and the self-reported sex, 15 participants due to high CRP and genes with few counts or high GC content influence, leaving 229 affected participants and 33 controls (Figure S1). Genes with less than 10 counts in at least 95% of the sample were removed, additionally, 1086 genes that were strongly influenced by the GC content were removed, leaving 12210 genes. The association of the batch variables with the first 10 principal components were assessed by ANOVA and visually inspected. The data was sequentially corrected for the GC content and a preparation batch variable with ComBat_seq from the sva package version 3.38.0 (Leek et al. 2021) and normalized with vst from DESeq2 version 1.30.1 (Love et al. 2021).

##### 1.4.3 Cell type deconvolution

The filtered RNA-seq count data, which was not batch-corrected, was converted to TPM data. This data was then deconvoluted using the granulator package version 1.6.0 (Pfister, Kuettel, and Ferrero 2023) in R version 4.2.0 (R Core Team 2023), applying the dtangle method. We utilized the leukocyte signature matrix (LM22) as a reference (Newman et al. 2015). All cell types with a standard variation of less than 0.5 were excluded, resulting in the 14 cell types B cells memory, B cells naive, Dendritic cells activated, Macrophages M0, Macrophages M1, Mast cells activated, Mast cells resting, Monocytes, NK cells activated, NK cells resting, T cells CD4 memory activated, T cells CD4 memory resting, T cells CD8 and T cells gamma delta.

##### 1.4.4 Multi-omics clustering and statistical analysis of cluster differences

Each variable of the normalized and batch corrected immune marker and RNA-seq data as well as age and BMI was standardized, and the distance between the participants within each layer was calculated with a Euclidean radial basis function. Here, we allowed for up to 20% missing values for a feature and 20% missing values for a participant. Subsequently, the non-negative matrix factorization-based clustering method, SUMO version 0.3.0 (Sienkiewicz et al. 2022) was applied using Python version 3.8.13 (Python Software Foundation 2022). This method executed 10,000 iterations for factorization for 2 to 5 clusters. We repeated the clustering 60 times on a 95% subsample (n=225) to calculate stability metrics for selecting the number of clusters. The optimal number of clusters was chosen based on the cophenetic correlation coefficient (bigger is better) and the proportion of ambiguously clustered pairs (smaller is better) as described in (Sienkiewicz et al. 2022). For detailed description of the clustering method, see (Sienkiewicz and Ratan 2022).

If not otherwise stated, we conducted the analyses using R version 4.0.2 (R Core Team 2023). We determined the variable importance based on the F-value from ANOVA tests, which tested the distribution of the variable of interest across different clusters. A higher variable importance indicates a more distinct separation between the clusters. For continuous variables not used in the clustering, we applied the Tukey Honest Significant Differences (TukeyHSD) test and reported adjusted p-values. We assessed the distribution of categorical variables, not included in the clustering, using Fisher’s exact test.

It is important to note that we did not report p-values for group differences for variables that were used in the clustering. Since the groups were defined based on these variables, differences were anticipated. Therefore, computing p-values in such a scenario would equate to “double dipping” (Kriegeskorte et al. 2009).

##### 1.4.5 Gene set analysis

For each participant, a score was calculated for every gene set using GSVA version 1.36.3. (Hänzelmann, Castelo, and Guinney 2013) based on the batch-corrected and normalized RNA-seq data. We included the biological process and molecular function GO term gene sets from version v2022.1 downloaded from MSigDB (Subramanian et al. 2005) that contain at least 10 genes and no more than 500 genes. The distribution of these gene set scores across clusters was then tested using an ANOVA, with F-values reported as variable importance. It is important to note that these F-values may be overoptimistic since the RNA-seq data was used in the clustering, hence a separation of gene sets is expected.

##### 1.4.6 Prediction of responder status

The gene expression data with accession number GSE45468 was downloaded with GEOquery version 2.58.0 (Davis 2023) and the baseline values extracted. The Illumina probe IDs were mapped to Entrez IDs with illuminaio version 0.32.0 (Baggerly et al. 2023) using the file HumanHT-12_V4_0_R2_15002873_B from <https://support.illumina.com/downloads/humanht-12_v3_product_files.html>, the Entrez IDs were checked for updates with rentrez version 1.2.3 (Winter, Chamberlain, and Guangchun 2020) and mapped to ENSEMBL IDs using the file hgnc_complete_set_2023-01-01.txt from <http://ftp.ebi.ac.uk/pub/databases/genenames/hgnc/archive/monthly/tsv/hgnc_complete_set_2023-01-01.txt>. If several probe IDs mapped to the same ENSEMBL ID, the first occurrence was kept. 8431 genes overlapped with our data and were used for the analysis. A lasso model was calculated with 4-fold cross validation using glmnet version 4.1-8 (Friedman et al. 2023). For predictions, lambda.min was used and the p-value was empirically calculated with 1000 models using random genes as comparison.

##### 1.4.7 Imaging analysis

Morphological differences were investigated at the global, regional (FreeSurfer) and voxel level. FreeSurfer v7.1.1 (Fischl 2012) based phenotypes were restricted to those with strong meta-analytical evidence for an effect in MDD as reported for 15 cortical thickness phenotypes (Schmaal et al. 2017) (Table S6), the bilateral hippocampal and lateral ventricle volume (Schmaal et al. 2016). Analysis of covariance (ANCOVA) was applied to the clusters covarying for intracranial volume, age and sex, and cluster effects were FDR corrected at 5%. For VBM of GM, a full factorial general linear model was estimated within the SPM12 framework, clusters collected at p_voxel_<0.001 and corrected for whole brain testing using FDR (significance at p_cluster.FDR_<0.05).

##### 1.4.8 Electrocardiography analysis

Raw data was preprocessed using the PhysioNet Cardiovascular Signal Toolbox (Vest et al. 2018) in Matlab version 2020b (The MathWorks Inc. 2020). Firstly, raw ECG data were converted to RR intervals through QRS detection. Next, the data were examined for possible confounders such as arrhythmia, ectopy, or low data quality. Flagged RR intervals were replaced with interpolated data. To ensure the integrity of the results and exclude potential biases caused by subjects who removed the ECG device before the end of the recording time, a visual quality control check was performed on all ECGs.

Time domain metrics were calculated for every minute using a sliding window of 5 minutes. The resulting variables included the highest and lowest normal-to-normal (NN) intervals (during sleep) in milliseconds, the overall variance in the distance between NN intervals, the standard deviation of NN intervals (SDNN) reflecting overall cardiac reactivity, and the root-mean-square between successive NN intervals (RMSSD) reflecting vagally mediated short-term variability in heart rate.

##### 1.4.9 Single-cell RNA-seq data analysis

Single cell PBMC RNA-seq data from 14 individuals with depression and other psychiatric disorders was downloaded from Gene Expression Omnibus with accession number GSE185714 and processed with scanpy version 1.9.3 (Wolf, Angerer, and Theis 2018) and Python version 3.9.17 (Python Software Foundation 2023) according to Schmid et al. 2021. The cell type labels B cells, CD4 T cells, CD8 T cells, CD14+ Monocytes, Dendritic cells, FCGR3A+ Monocytes, Megakaryocytes, NK cells and Plasma were also taken from Schmid et al. 2021.

### 2. Supplementary Results and Discussion

#### 2.1 Re-evaluating the initial clustering including cell types identified three clusters predominantly driven by RNA-seq data

Zinc Finger E-Box Binding Homeobox 2 (*ZEB2*), Phospholipase C Gamma 1 (*PLCG1*), Ras Homolog Family Member Q *(RHOQ),* Growth Arrest Specific 7 *(GAS7),* Enah/Vasp-Like *(EVL),* Adhesion Molecule With Ig Like Domain 1 *(AMIGO1)* and Long Intergenic Non-Protein Coding RNA 1550 *(LINC01550)* with VIs ranging from 85.3 to 68.8 were the essential variables to discriminate the clusters.

The cellular dynamics observed in our study were reflected in the gene expression patterns that most differentiated the reclusters. We noted elevated *ZEB2* expression in the reIDS and reHIRDS clusters consistent with increased immune cell counts in depression (Foley et al. 2022; Lynall et al. 2020), suggesting a link between immune activity and depression subtypes. *ZEB2*'s role in neural development and schizophrenia risk (Hegarty, Sullivan, and O’Keeffe 2015; Ripke et al. 2013; Khan et al. 2016) highlights the overlap between immune pathways and mental disorders. Additionally, *PLCG1*'s involvement in brain and immune signaling (Tubbs et al. 2020) further indicates the complexity of these interactions, emphasizing the need for further research into these molecular connections in mental disorders.

#### 2.2 Correlation between Immune markers and RNA-seq data

Notably, the correlation between the measured protein levels of VEGF-A and *VEGFA* gene expression was close to zero, in line with the other protein-gene pairs (Figure S10). In contrast, the correlation between the top differentiating genes was considerably higher, suggesting that they captured similar biological processes. The same was true for the top differentiating immune markers (Figure 4D-E). One possible reason for the poor correlation is post-transcriptional regulation that can control protein synthesis (Franks, Airoldi, and Slavov 2017), another the above discussed additional tissues that produce immune markers. In any case, this finding underscores the critical role of concurrently assessing protein levels and gene expression to fully understand the molecular dynamics.

### 3. Supplementary Figures

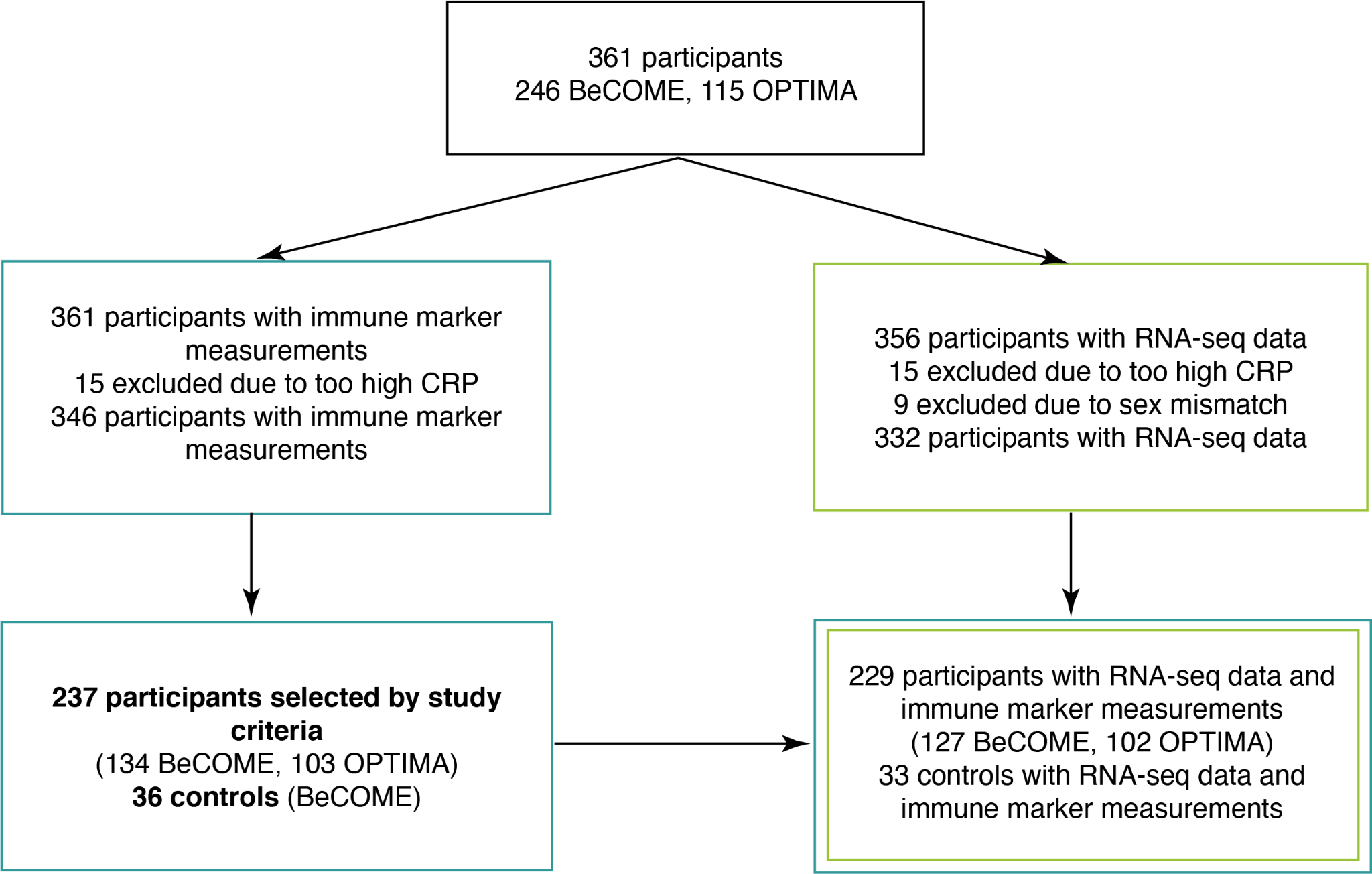

Figure S1: Overview about the sample size used. The study sample comprised 361 participants for which immune markers were measured. After exclusion of 15 participants due to too high CRP, 346 participants remained. Out of these, 237 participants were selected according to the study criteria and 36 healthy controls. Additionally, out of the 361 participants, 356 had RNA-seq data available. After exclusion of the 15 participants with too high CRP and 9 with a sex mismatch, 332 participants remained. Out of the selected 237 participants and 36 controls, 229 and 33 had both immune marker and RNA-seq data available. The immune marker data is available at Zenodo (<https://zenodo.org/doi/10.5281/zenodo.10698978>) and the RNA-seq data at GEO (accession number GSE260603).

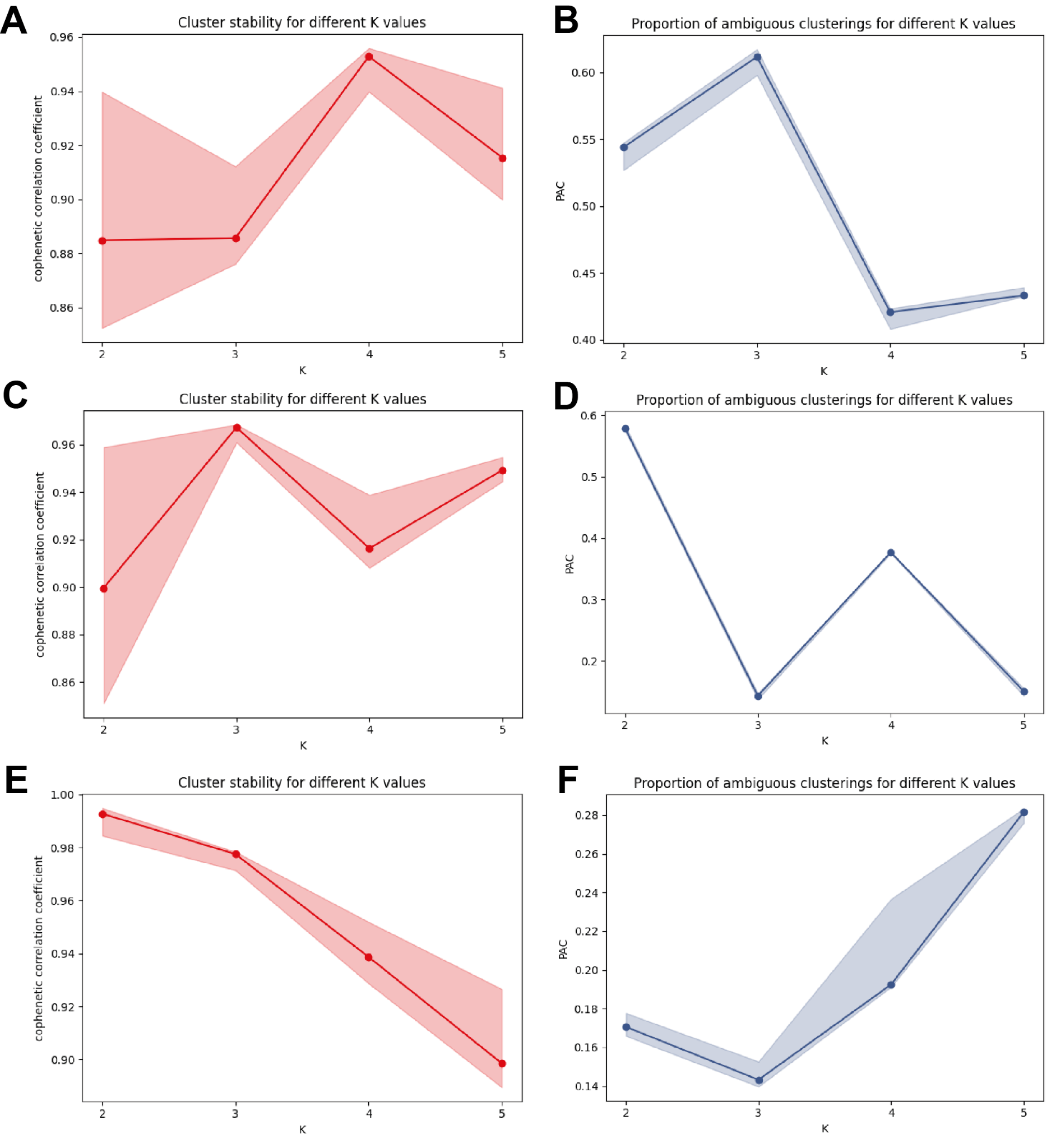

Figure S2: Line plots of the stability metrics for the SUMO clustering, for the cophenetic correlation coefficient a higher value is better and for the proportion of ambiguous clusterings a lower value is better. K denotes the number of clusters. (A) - (B) metrics for the initial clustering, (C) - (D) metrics for the clustering corrected for age, sex and BMI, (E) - (F) metrics for the re-clustering.

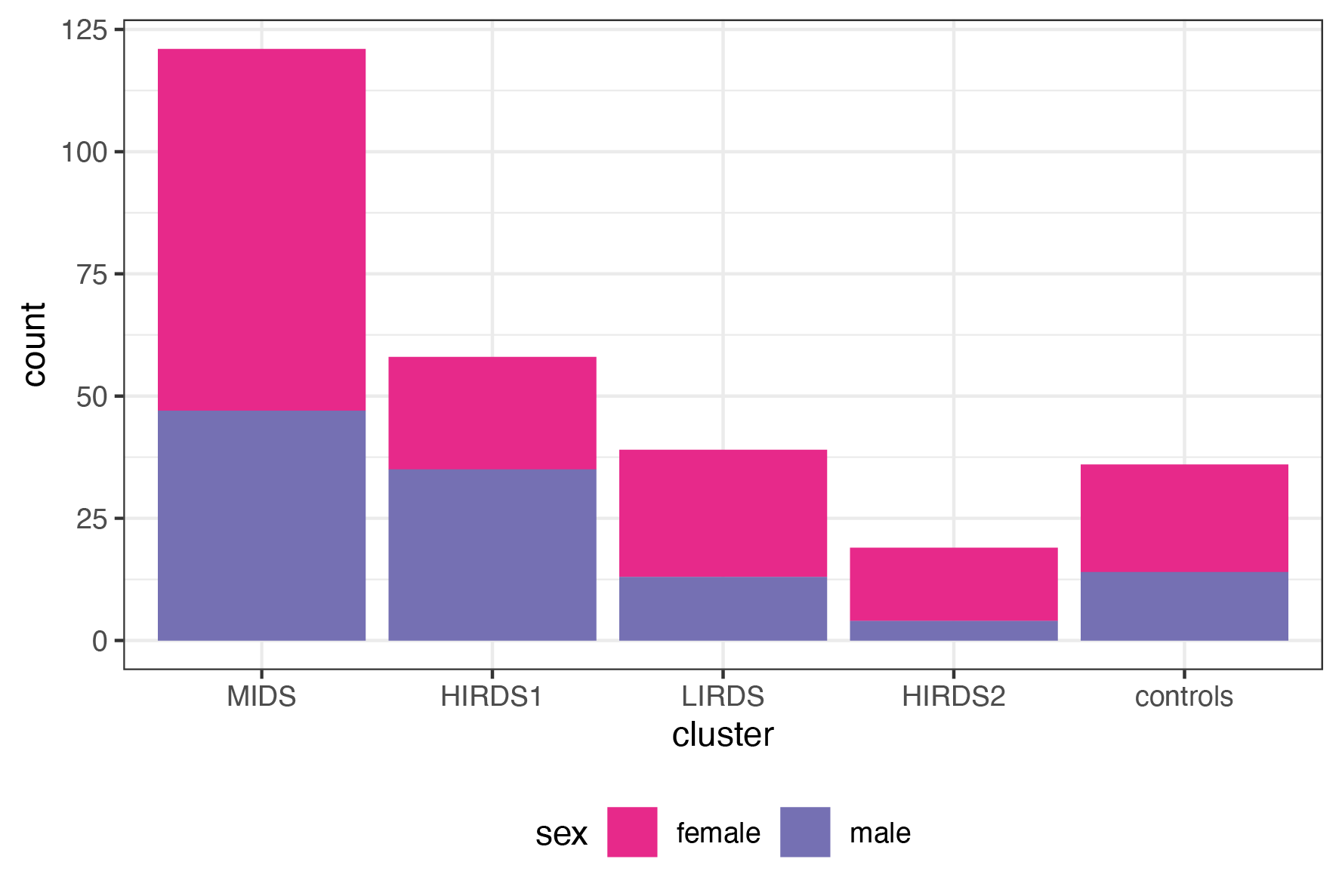
Figure S3: Sex distribution across the clusters of the initial clustering visualized as bar plots. N = 36 healthy controls not used in the clustering, MIDS = mild depression symptoms cluster, HIRDS = high immune-related depression symptoms cluster, LIRDS = low immune-related depression symptoms cluster.

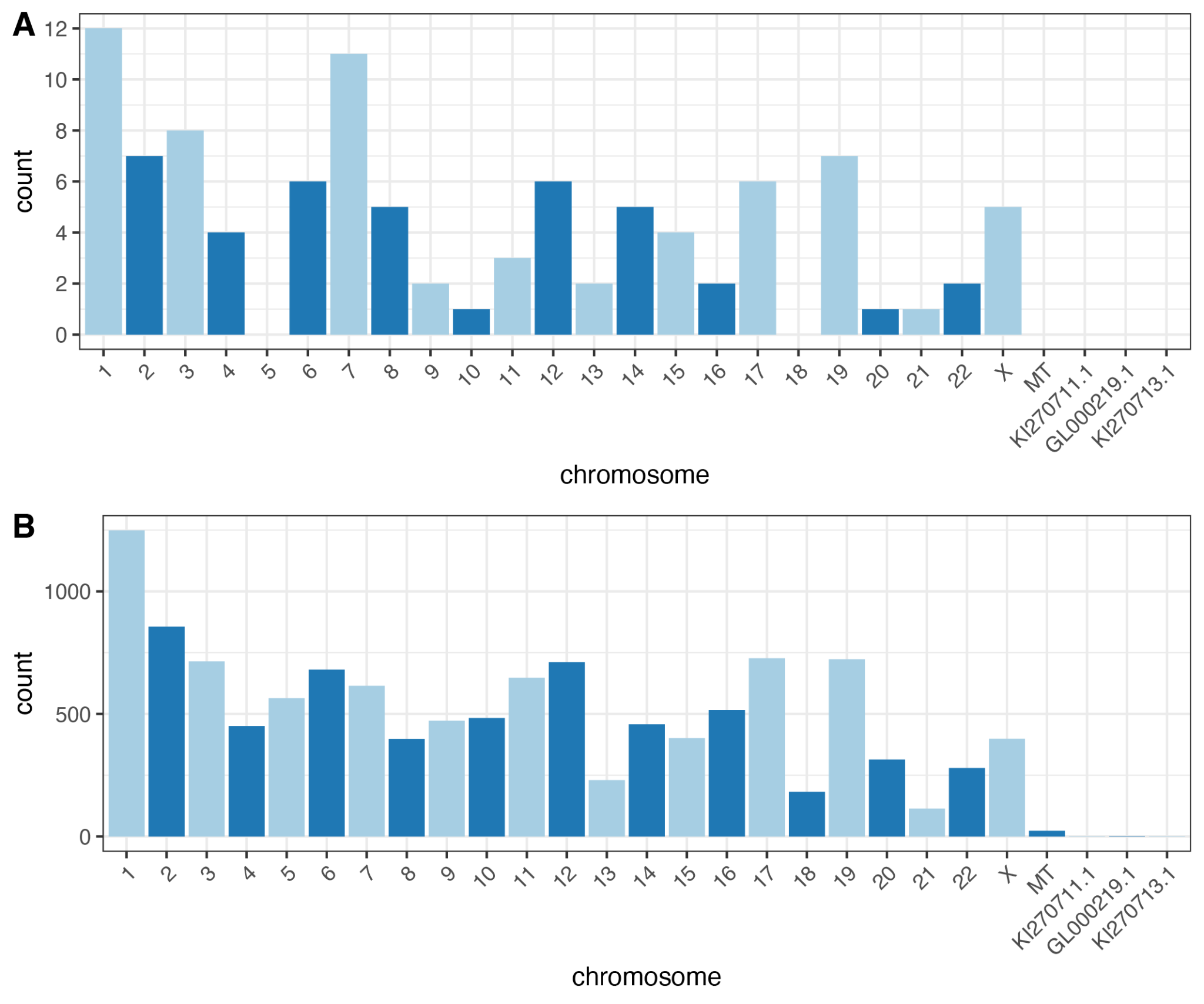
Figure S4: (A) Bar plot of the chromosomal distribution of the 100 genes with the highest variable importance of the initial clustering. (B) Bar plot of the chromosomal distribution of all 12210 genes mapped to GRCh38.p12.

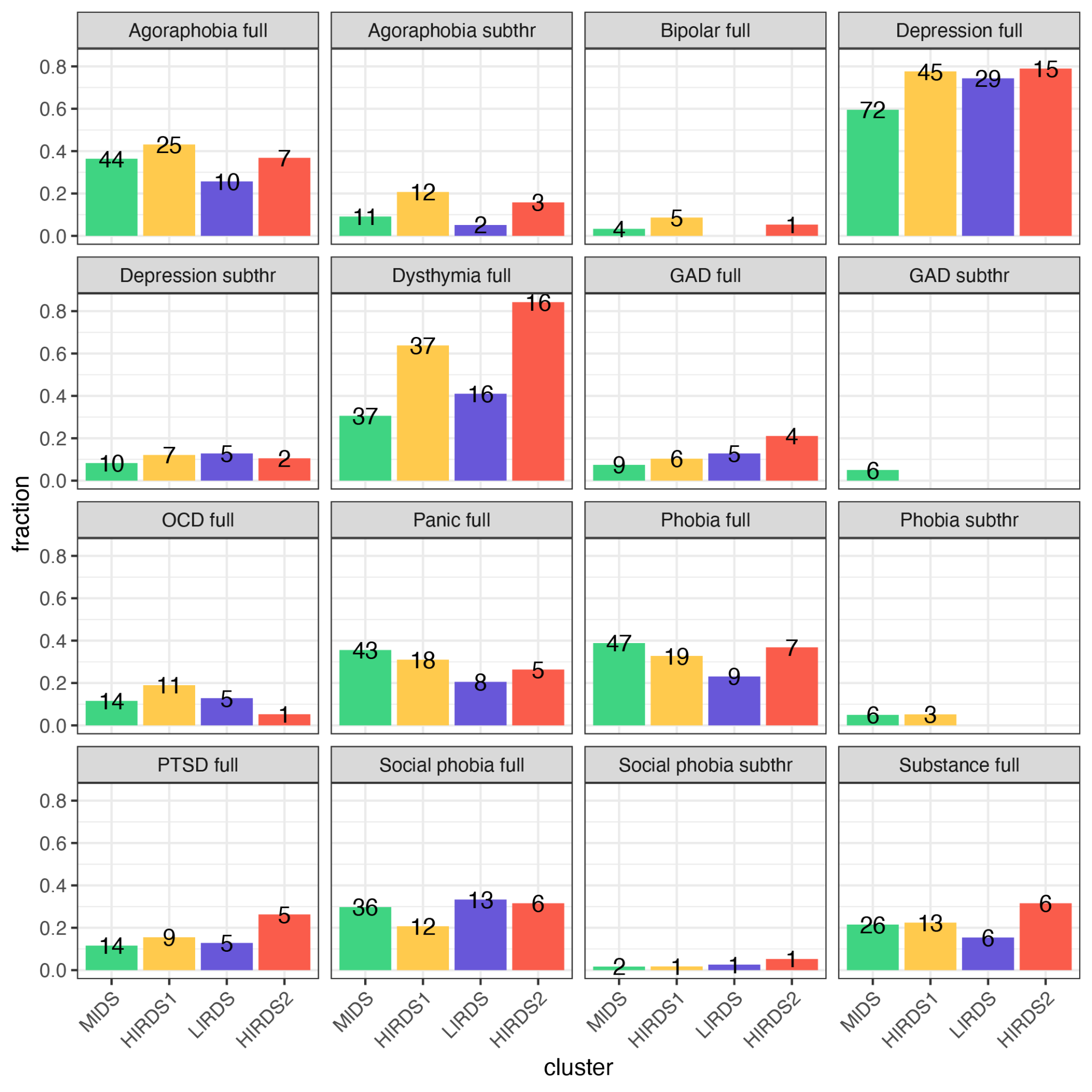

Figure S5: DIA-X/M-CIDI diagnostic categories distribution across the clusters of the initial clustering. The bar plots show the fraction while the numbers give the absolute number of participants. Full denotes that all criteria for a diagnosis in the DIA-X/M-CIDI were met, while subthr (subthreshold) denotes that only one criterion (e.g. impairment criterion) to the full-blown disorder was missing. Agoraphobia summarizes the ICD-10 codes F40.01, F40.00; Bipolar F31.8, F31.6, F31.5, F31.3, F31.2, F31.0, F30.0, F31.1; Depression F33.3, F33.2, F33.1, F33.0, F32.3, F32.2, F32.1, F32.0, Dysthymia F34.1; GAD = generalized anxiety disorder, F41.1; OCD = obsessive-compulsive disorder F42.8; Panic F41.0; Phobia F40.9, F40.2; PTSD = post-traumatic stress disorder F43.1; Social phobia F40.1; Substance F10.1, F10.2, F11.1, F11.2, F12.1, F12.2, F13.1, F13.2, F14.2, F15.1, F15.2, F16.2, F17.2, F18.1, F18.2, F19.1. MIDS = mild depression symptoms cluster, HIRDS = high immune-related depression symptoms cluster, LIRDS = low immune-related depression symptoms cluster.

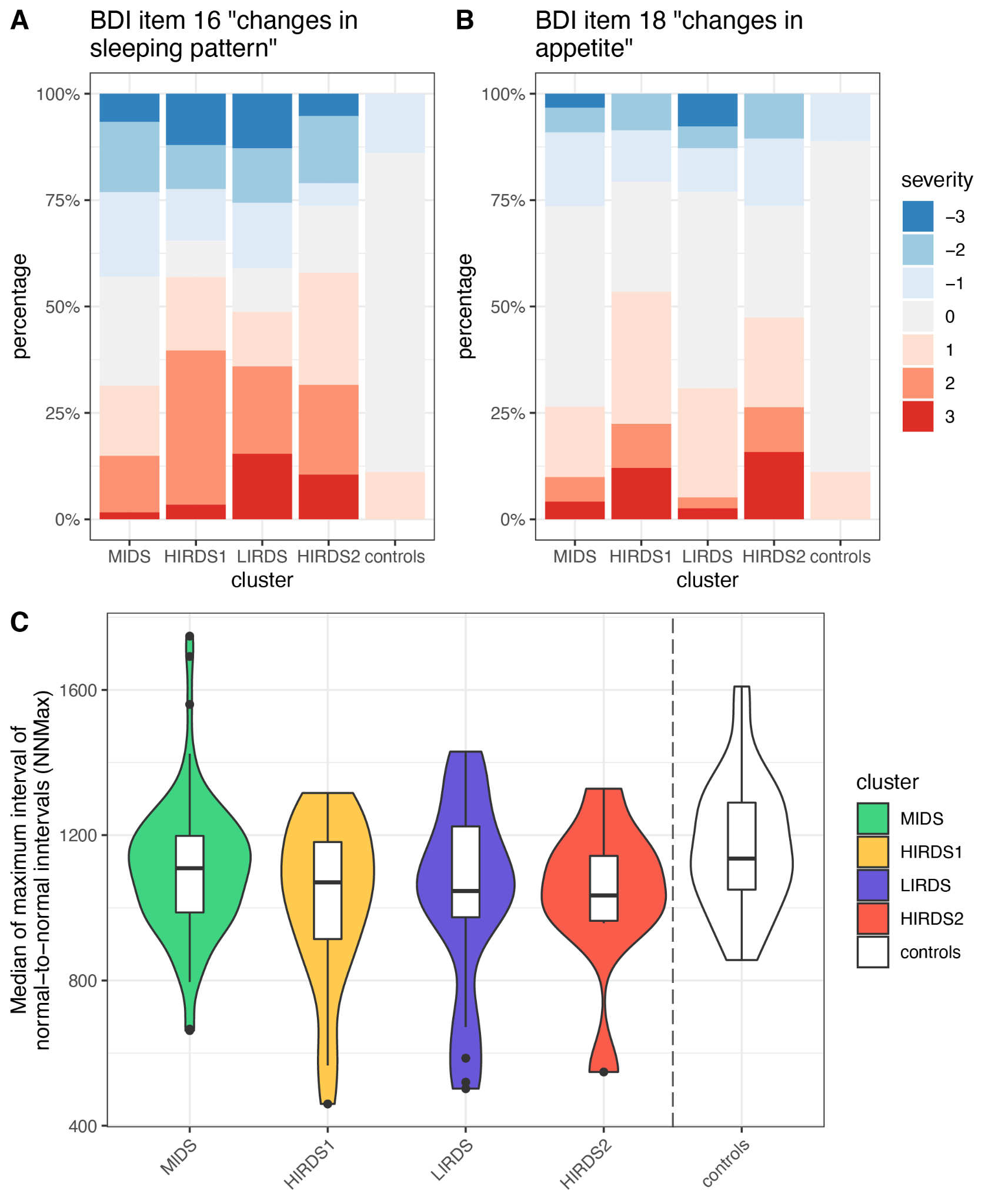

Figure S6: Bar plots showing the distribution of Beck Depression Inventory (BDI) item 16 “changes in sleeping pattern” (A) and item 18 “changes in appetite” (B) across the clusters of the initial clustering. Color intensities denote the severity with negative values (-1 to -3) indicating reduced sleep or appetite and positive values (1 to 3) indicating more sleep or appetite. The differences between the clusters were not significant. (C) Violin plots depicting the median of the maximum interval of NN intervals (NNMax), a measure for the heart rate for 149 participants. The observed differences between the clusters were not significant. N = 36 healthy controls not used in the clustering, MIDS = mild depression symptoms cluster, HIRDS = high immune-related depression symptoms cluster, LIRDS = low immune-related depression symptoms cluster.

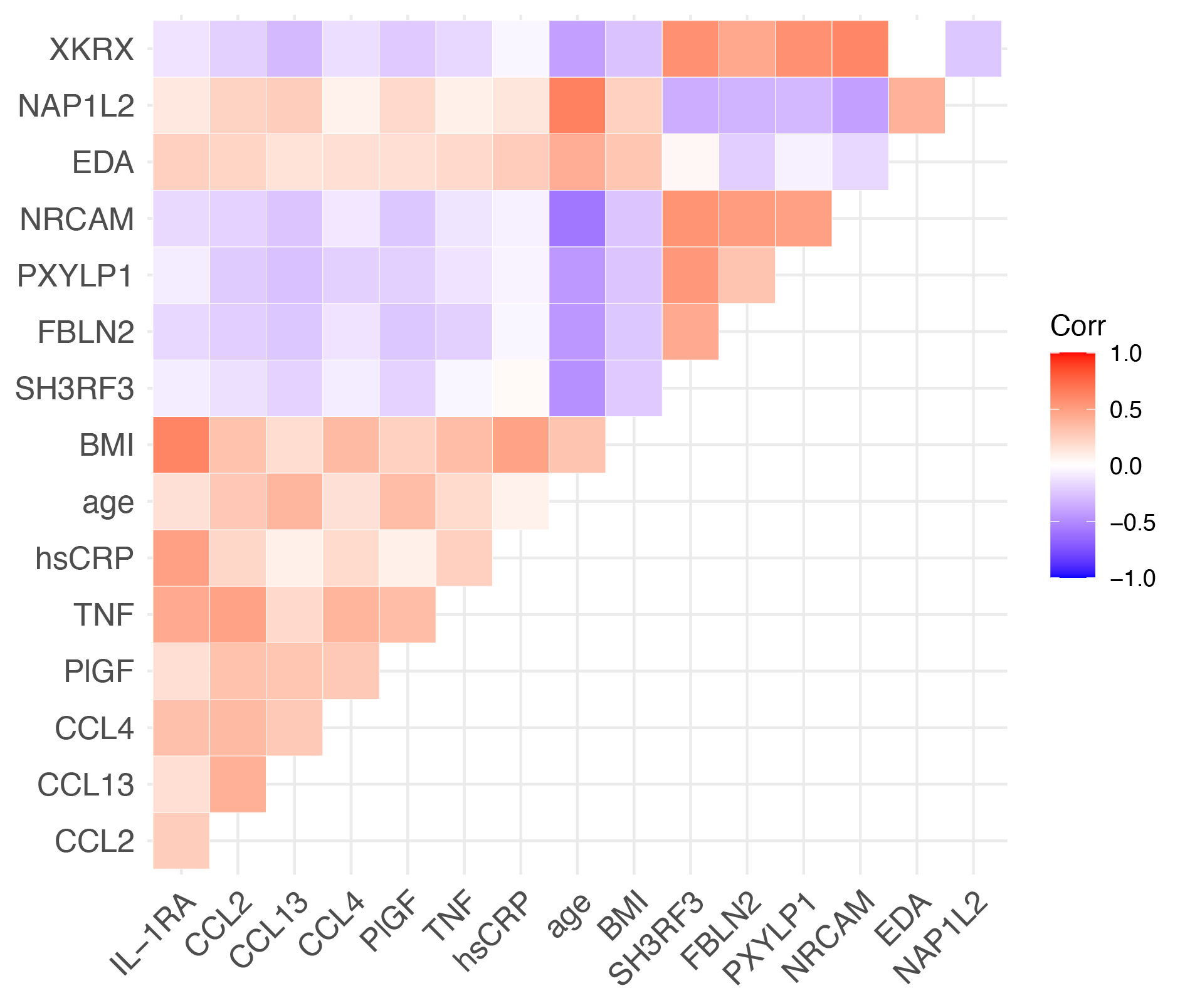

Figure S7: Pearson correlation of the immune markers, genes and BMI and age with the highest variable importance in the initial clustering.

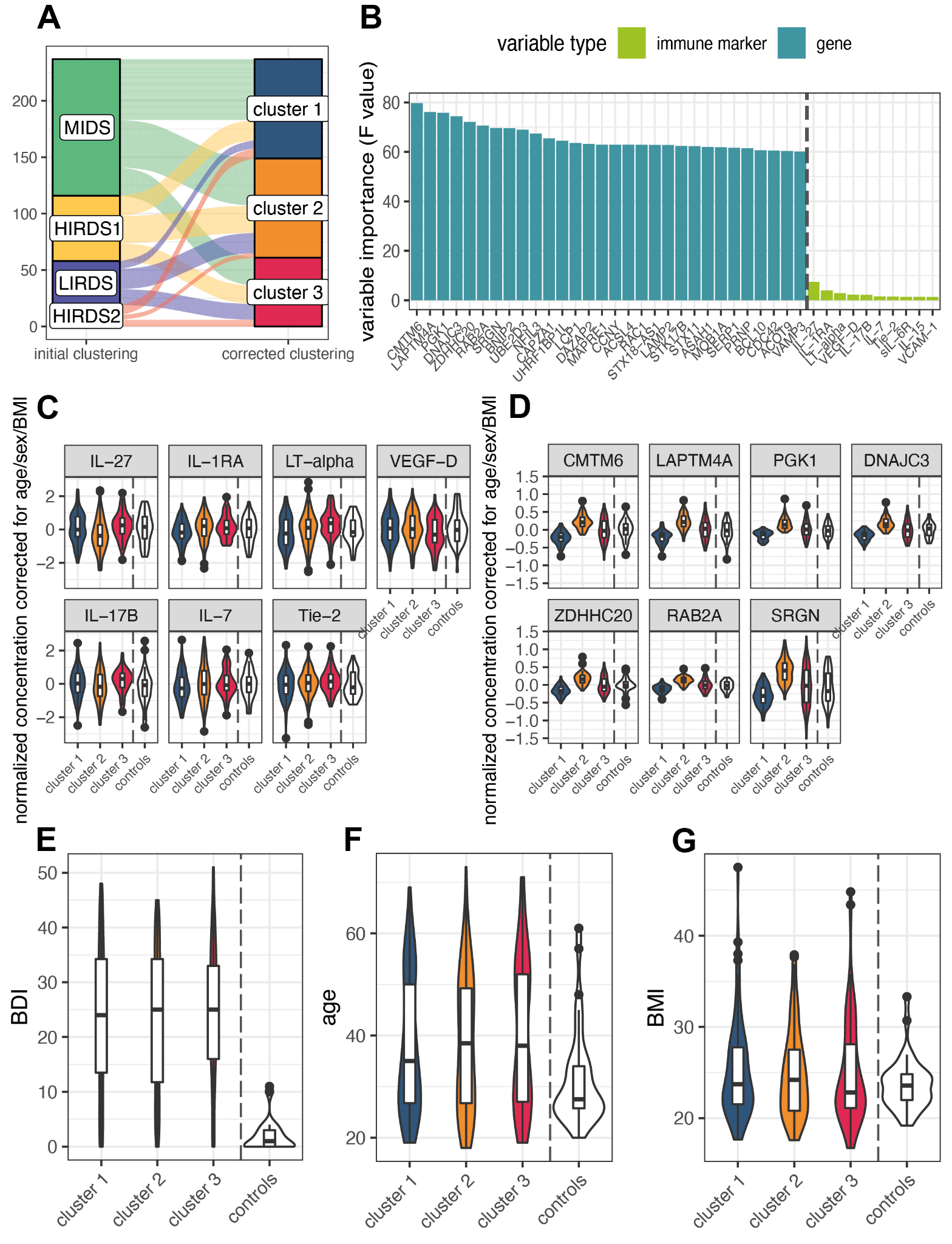

Figure S8: Clustering of immune markers and RNA-seq data corrected for age, sex and BMI. (A) Mapping of the participants from the initial clustering to the clustering corrected for age, sex and BMI. (B) Barplot showing the variable importance of the top 30 variables that most differentiate the clusters, along with the 10 most differentiating immune markers. (C) Violin plots showing the distribution of the seven most differentiating immune markers across the clusters. (D) Violin plots showing the distribution of the seven most differentiating genes across the clusters. (E) Violin plots showing the distribution of Beck Depression Inventory (BDI)-II sum score across the clusters, no significant differences. (F) Violin plots showing the distribution of age across the clusters, no significant differences. (G) Violin plots showing the distribution of BMI across the clusters, no significant differences. N = 36 healthy controls not used in the clustering.

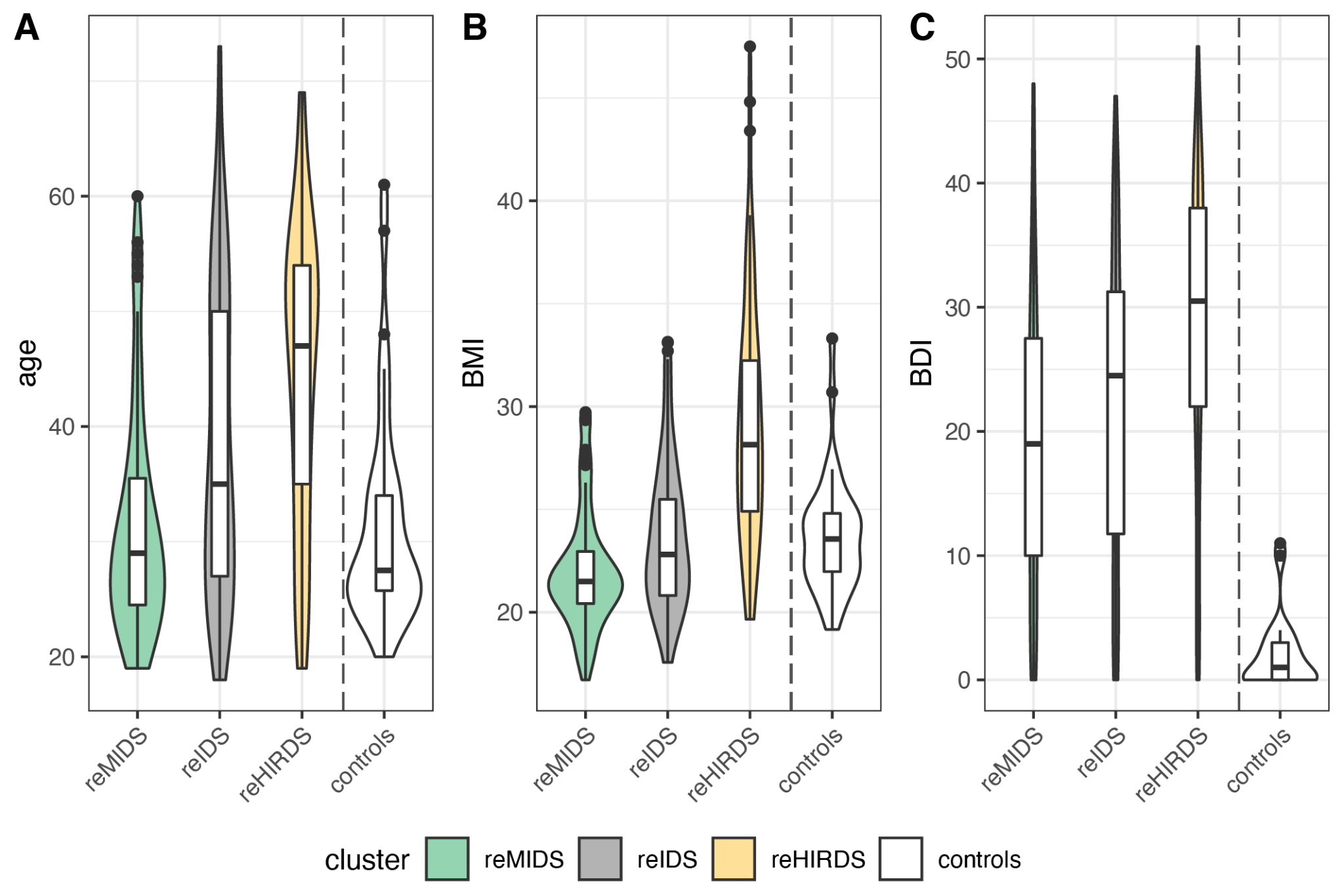

Figure S9: Re-clustering with age, BMI, cell types, immune markers and RNA-seq data. Violin plots depicting the distribution of (A) age, (B) BMI and (C) Beck Depression Inventory (BDI)-II sum score across the clusters. Significant differences (p < 0.05) for the BDI were observed when comparing the reHIRDS cluster to either the reMIDS or reIDS clusters. N = 36 healthy controls not used in the clustering, reMIDS = re mild depression symptoms cluster, reIDS = re intermediate depression symptoms cluster, reHIRDS = re high immune-related depression symptoms cluster.

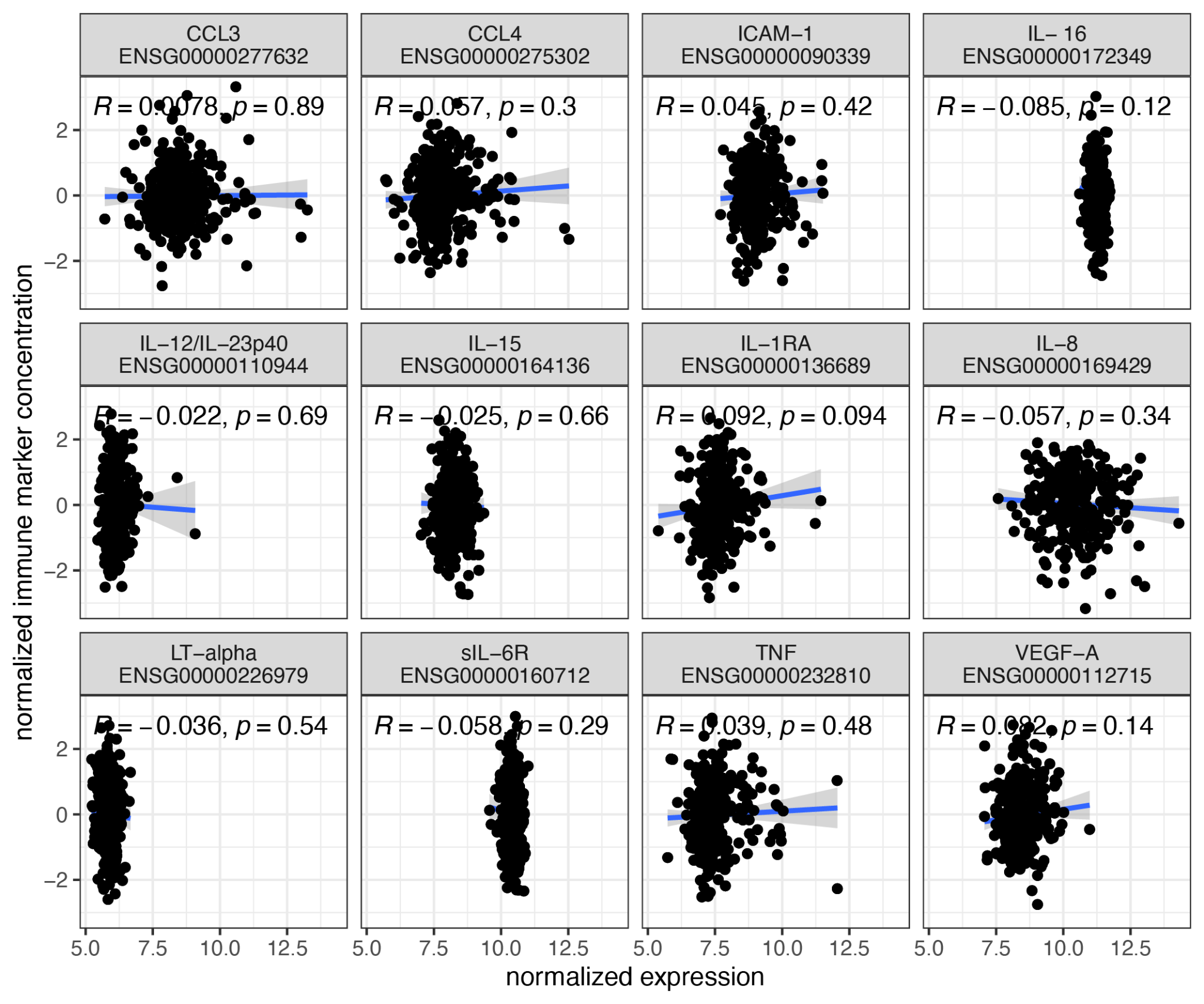

Figure S10: Scatter plots with the Pearson correlation of 12 immune markers for which both normalized protein concentrations and normalized gene expression data was available. The complete data set for which both RNA-seq and immune marker data was available was used, N = 332.

### 4. Supplementary Tables

Table S1: Somatic disease and medication information separated by status (cases used in the clustering and controls). N data available gives the number of individuals for which the information was available, the medication information was only available for the BeCOME cohort. N gives the number of individuals that took the medication or had the disease.

Table S2: Missingness information of the immune markers for the 237 individuals used in the clustering.

Table S3: Distribution of the BeCOME and OPTIMA participants across the clusters of the initial clustering, the clustering corrected for age/sex/BMI and the refined clustering refined including cell types. Also, the number of participants with available RNA-seq data, electrocardiography data (ECG) and structural imaging data is listed by cluster and study.

Table S4: Variable importance of the variables in the initial clustering calculated using the F-value from an ANOVA model with the clusters as independent variables. These variable importance values should not be interpreted as p-values, because the variables were used in the clustering themselves and therefore inflate the p-values.

Table S5: Genetic location of the top genes in the initial and revised clusterings with regard to GRCh38 based on the ENSEMBL data.

Table S6: Total brain volume (BV), gray matter (GM) and white matter (WM) compared between the MIDS, HIRDS1, LIRDS and HIRDS2 clusters, corrected for intracranial volume, age and sex with cluster sizes 89 (MIDS), 30 (HIRDS1), 22 (LIRDS) and 10 (HIRDS2). The pair-wise comparisons give nominal significance at p<0.05 denoted with * and a trend p-value (0.05 <= p < 0.10) denoted with +. SEM = standard error of the mean.

Table S7: List of tested FreeSurfer regional phenotypes (also see methods) and main effects of cluster comparison corrected for intracranial volume, age and sex between the MIDS (89), HIRDS1 (30), LIRDS (22) and HIRDS2 (10) clusters. The adjusted p-values are false discovery rate adjusted by the Benjamini-Hochberg procedure.

Table S8: Variable importance of the gene sets in the initial clustering calculated using the F-value from an ANOVA model with the clusters as independent variables. These variable importance values should not be interpreted as p-values, because the genes of the gene sets were used in the clustering.

Table S9: Variable importance of the variables in the clustering corrected for age, sex and BMI calculated using the F-value from an ANOVA model with the clusters as independent variables. These variable importance values should not be interpreted as p-values, because the variables were used in the clustering themselves and therefore inflate the p-values.

Table S10: Variable importance of the variables in the re-clustering calculated using the F-value from an ANOVA model with the clusters as independent variables. These variable importance values should not be interpreted as p-values, because the variables were used in the clustering themselves and therefore inflate the p-values.

Table S11: Variable importance of the gene sets in the re-clustering calculated using the F-value from an ANOVA model with the clusters as independent variables. These variable importance values should not be interpreted as p-values, because the genes of the gene sets were used in the clustering.

Hautzinger, Martin, Ferdinand Keller, and Christine Kühner. 2006. “BDI-II. Das Beck Depressions-Inventar II. Revision.” Pearson Deutschland.

Hegarty, Shane V., Aideen M. Sullivan, and Gerard W. O’Keeffe. 2015. “Zeb2: A Multifunctional Regulator of Nervous System Development.” *Progress in Neurobiology* 132 (September): 81–95. https://doi.org/10.1016/j.pneurobio.2015.07.001.

Khan, Raja Amjad Waheed, Jianhua Chen, Meng Wang, Zhiqiang Li, Jiawei Shen, Zujia Wen, Zhijian Song, et al. 2016. “A New Risk Locus in the ZEB2 Gene for Schizophrenia in the Han Chinese Population.” *Progress in Neuro-Psychopharmacology and Biological Psychiatry* 66 (April): 97–103. https://doi.org/10.1016/j.pnpbp.2015.12.001.

Kopf-Beck, Johannes, Petra Zimmermann, Samy Egli, Martin Rein, Nils Kappelmann, Julia Fietz, Jeanette Tamm, Katharina Rek, Susanne Lucae, and Anna-Katharine Brem. 2020. “Schema Therapy versus Cognitive Behavioral Therapy versus Individual Supportive Therapy for Depression in an Inpatient and Day Clinic Setting: Study Protocol of the OPTIMA-RCT.” *BMC Psychiatry* 20: 1–19.

Kriegeskorte, Nikolaus, W. Kyle Simmons, Patrick S. F. Bellgowan, and Chris I. Baker. 2009. “Circular Analysis in Systems Neuroscience: The Dangers of Double Dipping.” *Nature Neuroscience* 12 (5): 535–40. https://doi.org/10.1038/nn.2303.

Leek, Jeffrey T., W. Evan Johnson, Hilary S. Parker, Elana J. Fertig, Andrew E. Jaffe, Yuqing Zhang, John D. Storey, and Leonardo Collado Torres. 2021. “Sva: Surrogate Variable Analysis.” Bioconductor version: 3.12. https://doi.org/10.18129/B9.bioc.sva.

Liao, Yang, Gordon K. Smyth, and Wei Shi. 2014. “featureCounts: An Efficient General Purpose Program for Assigning Sequence Reads to Genomic Features.” *Bioinformatics* 30 (7): 923–30. https://doi.org/10.1093/bioinformatics/btt656.

Love, Michael, Constantin Ahlmann-Eltze, Kwame Forbes, Simon Anders, and Wolfgang Huber. 2021. “DESeq2: Differential Gene Expression Analysis Based on the Negative Binomial Distribution.” Bioconductor version: 3.12. https://doi.org/10.18129/B9.bioc.DESeq2.

Lynall, M. E., L. Turner, J. Bhatti, J. Cavanagh, P. de Boer, V. Mondelli, D. Jones, et al. 2020. “Peripheral Blood Cell-Stratified Subgroups of Inflamed Depression.” *Biol Psychiatry* 88 (2): 185–96. https://doi.org/10.1016/j.biopsych.2019.11.017.

Martin, Fergal J, M Ridwan Amode, Alisha Aneja, Olanrewaju Austine-Orimoloye, Andrey G Azov, If Barnes, Arne Becker, et al. 2023. “Ensembl 2023.” *Nucleic Acids Research* 51 (D1): D933–41. https://doi.org/10.1093/nar/gkac958.

Martin, Marcel. 2011. “Cutadapt Removes Adapter Sequences from High-Throughput Sequencing Reads.” *EMBnet.Journal* 17 (1): 10–12. https://doi.org/10.14806/ej.17.1.200.

Newman, Aaron M., Chih Long Liu, Michael R. Green, Andrew J. Gentles, Weiguo Feng, Yue Xu, Chuong D. Hoang, Maximilian Diehn, and Ash A. Alizadeh. 2015. “Robust Enumeration of Cell Subsets from Tissue Expression Profiles.” *Nature Methods* 12 (5): 453–57. https://doi.org/10.1038/nmeth.3337.

Pfister, Sabina, Vincent Kuettel, and Enrico Ferrero. 2023. “Granulator: Rapid Benchmarking of Methods for *in Silico* Deconvolution of Bulk RNA-Seq Data.” Bioconductor version: 3.16. https://doi.org/10.18129/B9.bioc.granulator.

Python Software Foundation. 2022. “Python.”

———. 2023. “Python.”

R Core Team. 2023. “R: A Language and Environent for Statistical Computing.” Vienna, Austria: R Foundation for Statistical Computing. https://www.R-project.org/.

Ripke, Stephan, Colm O’Dushlaine, Kimberly Chambert, Jennifer L. Moran, Anna K. Kähler, Susanne Akterin, Sarah E. Bergen, et al. 2013. “Genome-Wide Association Analysis Identifies 13 New Risk Loci for Schizophrenia.” *Nature Genetics* 45 (10): 1150–59. https://doi.org/10.1038/ng.2742.

Schmaal, L., D. P. Hibar, P. G. Sämann, G. B. Hall, B. T. Baune, N. Jahanshad, J. W. Cheung, et al. 2017. “Cortical Abnormalities in Adults and Adolescents with Major Depression Based on Brain Scans from 20 Cohorts Worldwide in the ENIGMA Major Depressive Disorder Working Group.” *Molecular Psychiatry* 22 (6): 900–909. https://doi.org/10.1038/mp.2016.60.

Schmaal, L., D. J. Veltman, T. G. M. van Erp, P. G. Sämann, T. Frodl, N. Jahanshad, E. Loehrer, et al. 2016. “Subcortical Brain Alterations in Major Depressive Disorder: Findings from the ENIGMA Major Depressive Disorder Working Group.” *Molecular Psychiatry* 21 (6): 806–12. https://doi.org/10.1038/mp.2015.69.

Schmid, Katharina T., Barbara Höllbacher, Cristiana Cruceanu, Anika Böttcher, Heiko Lickert, Elisabeth B. Binder, Fabian J. Theis, and Matthias Heinig. 2021. “scPower Accelerates and Optimizes the Design of Multi-Sample Single Cell Transcriptomic Studies.” *Nature Communications* 12 (1): 6625. https://doi.org/10.1038/s41467-021-26779-7.

Schmidtke, A, P Fleckenstein, W Moises, and H Beckmann. 1988. “Studies of the reliability and validity of the German version of the Montgomery-Asberg Depression Rating Scale (MADRS).” *Schweizer Archiv fur Neurologie und Psychiatrie (Zurich, Switzerland* 139 (2): 51–65.

Sienkiewicz, Karolina, Jinyu Chen, Ajay Chatrath, John T. Lawson, Nathan C. Sheffield, Louxin Zhang, and Aakrosh Ratan. 2022. “Detecting Molecular Subtypes from Multi-Omics Datasets Using SUMO.” *Cell Reports Methods*, 100152.

Sienkiewicz, Karolina, and Aakrosh Ratan. 2022. “Protocol for Integrative Subtyping of Lower-Grade Gliomas Using the SUMO Pipeline.” *STAR Protocols* 3 (1): 101110. https://doi.org/10.1016/j.xpro.2021.101110.

Smith, Tom, Andreas Heger, and Ian Sudbery. 2017. “UMI-Tools: Modeling Sequencing Errors in Unique Molecular Identifiers to Improve Quantification Accuracy.” *Genome Research* 27 (3): 491–99. https://doi.org/10.1101/gr.209601.116.

Subramanian, Aravind, Pablo Tamayo, Vamsi K. Mootha, Sayan Mukherjee, Benjamin L. Ebert, Michael A. Gillette, Amanda Paulovich, et al. 2005. “Gene Set Enrichment Analysis: A Knowledge-Based Approach for Interpreting Genome-Wide Expression Profiles.” *Proceedings of the National Academy of Sciences* 102 (43): 15545–50. https://doi.org/10.1073/pnas.0506580102.

The MathWorks Inc. 2020. “MATLAB Version: R2020b.” Natick, Massachusetts, United States: The MathWorks Inc. https://www.mathworks.com.

Tubbs, Justin D., Jiahong Ding, Larry Baum, and Pak C. Sham. 2020. “Immune Dysregulation in Depression: Evidence from Genome-Wide Association.” *Brain, Behavior, & Immunity - Health* 7 (August): 100108. https://doi.org/10.1016/j.bbih.2020.100108.

Vest, Adriana N., Giulia Da Poian, Qiao Li, Chengyu Liu, Shamim Nemati, Amit J. Shah, and Gari D. Clifford. 2018. “An Open Source Benchmarked Toolbox for Cardiovascular Waveform and Interval Analysis.” *Physiological Measurement* 39 (10): 105004. https://doi.org/10.1088/1361-6579/aae021.

Winter, David, Scott Chamberlain, and Han Guangchun. 2020. “Rentrez: ‘Entrez’ in R.” https://cran.r-project.org/web/packages/rentrez/index.html.

Wittchen, H.-U., E. Beloch, E. Garzcynski, A. Holly, G. Lachner, A. Perkonigg, E.-M. Pfütze, et al. 1995. “Münchener Composite International Diagnostic Interview (M-CIDI) (Version 2.2 / 2 / 95).” Max-Planck-Institut für Psychiatrie, Klinisches Institut, München.

Wittchen, H.-U., and H. Pfister. 1997. “DIA-X-Interviews: Manual für Screening-Verfahren und Interview; Interviewheft.” https://hdl.handle.net/11858/00-001M-0000-000E-AAAF-C.

Wolf, F. Alexander, Philipp Angerer, and Fabian J. Theis. 2018. “SCANPY: Large-Scale Single-Cell Gene Expression Data Analysis.” *Genome Biology* 19 (1): 15. https://doi.org/10.1186/s13059-017-1382-0.
